# ‘Leading from the front’ implementation strategies increase the success of influenza vaccination drives among healthcare workers: A reanalysis of Systematic Review evidence using Intervention Component Analysis (ICA) and Qualitative Comparative Analysis (QCA)

**DOI:** 10.1101/2021.02.10.21251475

**Authors:** Katy Sutcliffe, Dylan Kneale, James Thomas

**Author notes:** Corresponding author Dr Katy Sutcliffe.

## Abstract

**Background:** Seasonal influenza vaccination of healthcare workers (HCW) is widely recommended to protect staff and patients. A previous systematic review examined interventions to encourage uptake finding that hard mandates, such as loss of employment for non-vaccination, were more effective than soft mandates, such as signing a declination form, or other interventions such as incentives. Despite these overarching patterns the authors of the review concluded that ‘substantial heterogeneity’ remained requiring further analysis. This paper reanalyses the evidence using Intervention Component Analysis (ICA) and Qualitative Comparative Analysis (QCA) to examine whether the strategies used to implement interventions explain the residual heterogeneity.

**Methods:** We used ICA to extract implementation features and trialist’s reflections on what underpinned the success of the intervention they evaluated. The ICA findings then informed and structured two QCA analyses to systematically analyse associations between implementation features and intervention outcomes. Analysis 1 examined hard mandate studies. Analysis 2 examined soft mandates and other interventions.

**Results:** In Analysis 1 ICA revealed the significance of ‘leading from the front’ rather than ‘top-down’ implementation of hard mandates. Four key features underpinned this: providing education prior to implementation; two-way engagement so HCW can voice concerns prior to implementation; previous use of other strategies so that institutions ‘don’t-go-in-cold’ with hard-mandates; and support from institutional leadership. QCA revealed that either of two configurations were associated with greater success of hard mandates. The first involves two-way engagement, leadership support and a ‘don’t-go-in-cold’ approach. The second involves leadership support, education and a ‘don’t-go-in-cold’ approach. Reapplying the ‘leading from the front’ theory in Analysis 2 revealed similar patterns.

**Conclusions:** Regardless of intervention type a ‘leading from the front’ approach to implementation will likely enhance intervention success. While the results pertain to flu vaccination among HCWs, the components identified here may be relevant to public health campaigns regarding COVID-19 vaccination.

## Introduction

Seasonal influenza can have dire consequences for individuals, particularly for vulnerable groups such as children, older people and those with pre-existing health problems [1]. Outbreaks can also place significant strain on health services. This can result from both an increased number of patients, and from a reduced number of available healthcare workers (HCW) as their role puts them at high risk of infection due to close contact with the virus [2]. In order to protect themselves and their patients HCWs involved in direct patient care are encouraged to receive an influenza vaccine [3]. Whilst evidence shows influenza vaccine to be safe, effective, and to decrease mortality in patients [4] a key challenge is poor vaccine uptake. In the 2018-2019 season in England 70% of frontline HCWs were vaccinated, which represents a year-on-year increase, but is short of the national target of 75% [3]. Vaccine hesitancy has been increasing in recent years [5, 6] and the COVID-19 pandemic has highlighted the urgency of understanding how to address it [7], particularly among HCWs to ensure their wellbeing as well as to esnure the delivery of safe, efficient and effective healthcare services [8].

A comprehensive systematic review [9], which was recently updated [10], found that various interventions to encourage uptake can increase rates of vaccination among HCW. The review examined both voluntary programmes (such as incentives, media campaigns or education programmes) and policies which make vaccination mandatory for HCWs. Meta-analysis was used to quantify the effects of the various approaches in the original review. The findings demonstrated that among the intervention strategies examined, ‘hard’ mandates such as loss of employment for non-vaccination were by far the most effective (RR_unvac_ (risk ratio of being unvaccinated) = 0.18, 95% CI: 0.08–0.45). This was followed by ‘soft’ mandates such as requiring staff to sign a declination form, increasing access (i.e. making it easier for staff to receive the vaccination) (RR_unvac_ = 0.64, 95% CI: 0.45–0.92) and increasing awareness (e.g. through media campaigns) (RR_unvac_ = 0.83, 95% CI: 0.71– 0.97). The pooled findings for incentives did not quite reach statistical significance (RR_unvac_ = 0.89, 95% CI: 0.77–1.03) and pooled findings for educational interventions showed no evidence of an effect (RR_unvac_ = 0.96, 95% CI: 0.84–1.10).

Whilst these pooled findings about the pooled effects of interventions within broad categories is a useful step in understanding how best to address the issue of vaccination uptake in HCW, vital knowledge about exactly what to implement and how is lacking. The authors identified ‘substantial heterogeneity’ in the findings ([9] p.66) and acknowledged that this may be due to a number of factors including: the HCW populations studied; the clinical setting; the country; the specific components of each intervention and the way these were implemented in each study. For example, the exact nature of ‘hard-mandates’ varied considerably; some required mask use for unvaccinated HCW whilst others prohibited patient contact and yet others resulted in termination of employment.

Uptake of the review findings may therefore be hindered by a lack of information about the specific features and implementation methods of successful strategies [11, 12]. In addition, ethical concerns about the use of hard mandates suggest a more holistic understanding of such strategies is warranted [13]. The aim of this project was to reanalyse the trials using an alternative analytical technique – qualitative comparative analysis (QCA). QCA – originally developed in political science [14] – has recently been employed in systematic reviews [15, 16]. The technique seeks to uncover the causal mechanisms and key features of an intervention. QCA is a ‘case’ rather than a ‘variable’ oriented approach. A ‘case’ in QCA essentially refers to a study - both its features and the context in which it was implemented. And the ‘case’ oriented approach requires a deep and holistic understanding of each case. Another key feature of QCA is that it uses set theory. QCA makes systematic comparisons between cases based on their outcomes – i.e. comparing the characteristics of a set (i.e. a group) of effective interventions to those of a set of ineffective interventions. QCA seeks to identify the degree of overlap between these outcome sets and sets of interventions with similar characteristics. This approach enables an analysis that, unlike statistical approaches, can operate with relatively small numbers of studies and a large number of variables (which are referred to as ‘conditions’ in QCA). Lastly QCA is an abductive approach. Unlike the deductive approach of meta-analysis in which a hypothesis is posed and then tested, the abductive approach involves starting with an observed outcome (in this case rates of vaccination uptake) and working backwards to identify the simplest and most likely explanation for the observed outcome. Because the abductive approach yields a plausible explanation but is not able to conclusively verify it, it is far less secure than a deductive approach. As such a key requirement is that the analysis is underpinned by theory.

The high-level findings of the Lytras et al. review about the success of hard-mandates suggest the validity of a ‘sticks are better than carrots’ intervention theory. However, since not all hard-mandate (or soft-mandate) interventions achieved similar rates of success we needed to look beyond the overt intervention theory and to focus on ‘on-the-ground’ implementation and context. Intervention Component Analysis (ICA) is a methodological approach which seeks to ‘bridge the gap’ between evidence of intervention effectiveness and practical implementation of interventions [17]. More specifically, ICA seeks to generate an ‘experienced-based’ understanding of intervention mechanisms by tapping into trialist’s informal reflections about how the interventions they evaluated worked ‘on the ground’. ICA uses qualitative data analysis techniques and draws on informal evidence – often reported in the discussion section of published trial reports – about what trialists’ felt to led to the success of an intervention or what inhibited its success. Of course, there are potential limitations to drawing on informal data of this kind. However, ICA offers a systematic process through which experience-based theoretical explanations of intervention mechanisms can be developed, and which can then be tested using more formal analytical techniques such as QCA. In addition, given that (too) many outcome evaluations fail to be accompanied by a process evaluation, which could provide richer data on intervention mechanisms and fidelity to intervention protocols, ICA provides a framework for incorporating additional data on intervention processes and components. ICA and QCA were paired in a previous project to successfully identify critical intervention mechanisms [18].

The overarching aim of this research was to support hospitals to implement effective vaccination uptake strategies by identifying the critical features and implementation methods of successful strategies. In addition, by exploring how vaccination uptake strategies work, we hoped to provide some insights that might assist with global drives to vaccinate against COVID-19.

## Materials and methods

The research involved a reanalysis of the trials included in the Lytras et al. 2016 review [9] and from the Lorenc et al. 2018 update [10]. Ethical approval was not obtained since the analysis involved only published data already in the public domain. There are no reporting guidelines for reanalyses of systematic reviews, although guidance for QCA studies is being developed [19] and we have sought to provide a detailed and transparent account of the work such that it could be replicated.

Our initial hypothesis was that the mechanisms differentiating the more successful from the less successful hard-mandate interventions would differ from the mechanisms differentiating the more successful of the soft-mandate and other interventions from those that were less successful. Thus, we conducted two separate analyses. Analysis 1 explored which intervention and implementation features were associated with greater effectiveness among the hard-mandate interventions, and Analysis 2 explored which features were associated with greater effectiveness among the soft-mandate and other interventions. We completed all of the QCA stages for Analysis 1 before repeating the process for Analysis 2.

### QCA stage 0: Selection of cases and determining outcome sets

For Analysis 1 we selected all eight of the hard mandate cases [20-26] included in the original review [9] (note: two hard mandate cases were evaluated in the Ksienski 2014 study), and the three additional hard mandate cases [27-29] identified in the update [10]. For Analysis 2 there was a much greater number of non-hard mandate cases (45 cases from the review and 12 from the update) so we were able to purposively select the cases with maximum variation in outcomes, i.e. the 10 most effective non-hard mandate cases [30-37] and the 10 least effective ones [38-44]. (Note: A total of six papers reported the 10 least effective soft mandate / other cases; two cases were reported in each of the following three papers Dey et al. 2001, Doratotaj et al. 2008 and Zimmerman et al. 2009.) By excluding the moderately effective non-hard mandate cases we filtered out ‘noise’ which might obscure differences between the most effective and least effective. Effectiveness was determined as per the original Lytras review in terms of the Relative Risk of remaining unvaccinated after the intervention (RR_unvac_); values of RR_unvac_ < 1 suggest that the intervention is effective in reducing the number of unvaccinated HCWs. For Analysis 2 we used crisp outcome sets, in which cases are full members of a set of ‘most effective’ cases or full members of a set of ‘least effective cases’. We ranked the cases according to their RR_unvac_ value; the 10 in the most effective set had values ranging from 0.06 to 0.59, the 10 in the least effective set had values ranging from 0.95 to 0.99. Since we included the full range of outcomes for Analysis 1 (i.e. we did not exclude moderately effective cases as we did for Analysis 2) we created fuzzy outcome sets, where studies could be partial members of sets. A fully successful outcome set (coded as 1) comprised of four cases with RR_unvac_ values between 0.01 and 0.14. A mostly successful outcome set (coded as 0.66) comprised of four cases with RR_unvac_ values between 0.15 and 0.29. A mostly unsuccessful outcome set (coded as 0.33) comprised of two cases with RR_unvac_ values between 0.30 and the least effective in the set (0.57).

### QCA stage 1: Identification of conditions using ICA and building the data table

Once we had selected our cases and determined our outcome sets we read and re-read the papers reporting the 11 hard-mandate cases to generate a deep knowledge for Analysis 1. After the familiarisation exercise two authors (KS and DK) independently extracted information about the nature of the hard-mandate interventions to create a data table with cases represented in rows and conditions represented in columns (see supporting information). Initial work focused on the intervention descriptions as provided by the authors – for example we captured data on the nature of hard mandates such as whether it resulted in loss of employment or not, whether there were stigmatising markers of non-identification and whether any ‘declination’ procedures were particularly onerous or not. However, limiting our data collection to the intervention descriptions alone proved unfruitful for identifying features that distinguished between the most and least successful interventions. Thus, we decided to focus on implementation and to employ ICA to extract information from the discussion section. Specifically, we used inductive qualitative analysis techniques to code authors’ perceptions about the factors that acted as facilitators of or barriers to success. ICA revealed four implementation features that were commonly described by authors as underpinning the success of hard mandate interventions: **Education** (reported in 5 cases) for example providing information sessions prior to mandate implementation; **two-way engagement** (reported in 2 cases) i.e. opportunities for HCW to raise concerns; **‘don’t go in cold’** (reported in 5 cases) i.e. efforts in previous years to encourage vaccination uptake; and **leadership support** (reported in 6 cases) i.e. involvement and endorsement from senior leaders in the institution. Box 1, below provides example statements from authors regarding the importance of these implementation features. Before proceeding to the next stages of QCA analysis the quality of the data was evaluated, including checks for ‘collinearity’ of conditions and rarity of conditions.

**Box 1: Example author statements about factors perceived as vital to successful hard-mandate implementation**

**Education:** “*Key factors that supported the success of the program included consistent communication emphasizing patient safety and quality of care*.” (Babcock et al. 2010)

**Two-way engagement:** *“Continued stakeholder engagement is required to ensure that the decision-making process is collaborative and the Policy is not viewed as punitive*.*”* (Ksienski 2014)

**Don’t go in cold:** *“Sequential expansion of the program over several years was a key element to the success*.*”* (Frenzel et al. 2016)

**Leadership support:** *“Without a strong endorsement from the CEO, president, and governing board, it is unlikely that the program would have been successful*.*”* (Rakita et al. 2010)

We returned to the theoretical literature to see if existing theories reflecting our emergent findings could help to consolidate our thinking. This process identified the theoretical concept of ‘leading from the front’ as opposed to a ‘top-down’ or ‘authoritarian’ approach to leadership with the key underpinning principle being that organisations should aim to ‘bring people with you’. The concept draws on literature on transformational leadership which emphasises communication, listening, modelling and leadership commitment [45].

The same steps were taken for Analysis 2; however as we had assumed a different mechanism would underpin the non-hard mandate studies we did not initially extract the same conditions as identified in the ICA for Analysis 1. Initial work for Analysis 2 was based on a ‘dark logic’ approach [46]. Since the non-hard mandate interventions were found to be broadly less effective than hard-mandate interventions we considered whether we might identify harmful or ineffective mechanisms that undermined the approach. However, this analytical plan proved unfruitful. So we decided to see if the same conditions and the ‘leading from the front’ theory might also explain the variation in outcomes among the soft-mandate and other interventions.

### QCA stage 2: Constructing Truth Tables

In QCA stage 2 a Truth table, the key analytic device of QCA, is created. The Truth Table moves the focus from individual cases to groups of cases sharing the same outcomes ‘outcome sets’ (as described above) and from individual conditions to sets of studies with particular combinations or “configurations” of conditions. The Truth Tables for analyses 1 (Table 1) and 2 (Table 3) are presented below.

**Table 1:**
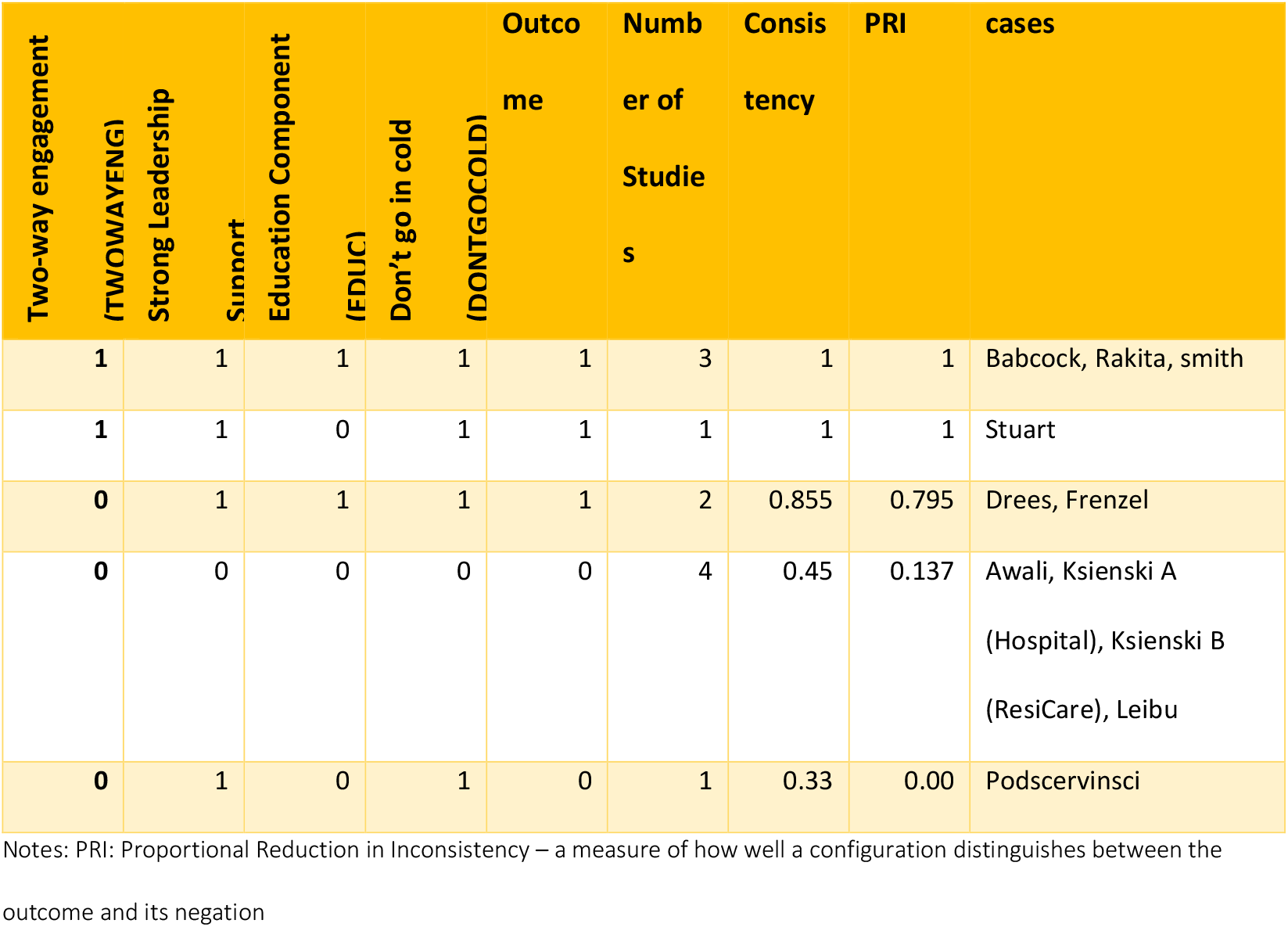
**Truth Table for Hard Mandate QCA (n=11 cases)**

### QCA stage 3: Checking the quality of the Truth Tables

The first check of each Truth Table involved assessing the degree to which a consistent pattern of association between the configurations and the outcome sets is evident. For example, if all cases involving all four conditions in the theory (education, two-way engagement, a ‘don’t go in cold’ approach and leadership support) are also all cases that are fully part of the successful outcome set and none are cases in the unsuccessful outcome set, that would show a perfect consistency score, indicated with a ‘1’, for that row of the Truth Table. Conversely, if all cases in which none of the four conditions were present were also all cases in the unsuccessful outcome set, this would also show perfect consistency and be indicated by a ‘0’. Some level of inconsistency is permitted and even expected with fuzzy-set QCA – but patterns of association should be evident, and inconsistency explored for potential deviant cases; for crisp-set QCA, inconsistency is not expected and needs to be resolved or explained. The second check we performed was to assess coverage, i.e. whether configurations are supported by multiple cases. It is expected that there will be several paths to a given outcome, and so the coverage offered by any given configuration may only be one or a small number of cases. However, where multiple cases support a configuration - it helps us to understand the relevance or importance of different configurations, and reduces the possibility that the resulting QCA solution becomes an explanation of individual cases. A third check examined whether there was a reasonable spread of cases across the 16 possible configurations in each of our truth tables. Having evidence for a range of possible configurations helps us to interpret and refine our causal theory. Final checks included (i) examining for deviant cases consistency [47] - those cases with values above 0.5 for the condition configuration and below 0.5 for the outcome (inconsistencies); and (ii) examination of counterintuitive findings – e.g. if cases with all conditions specified in our underlying theory were associated with unsuccessful outcomes – indicating that our theory does not play out in practice. As the Truth Tables below illustrate, we found satisfactory results for each of the above checks.

### QCA stage 4: Boolean minimization to identify the simplest expression of configurations

We used Boolean minimisation to identify simplified configurations with coverage of as many of the cases in the successful outcome set as possible and with high consistency, generating what is known in QCA parlance as a ‘complex solution’.

### QCA stage 5: Consideration of “logical remainders”

In this stage possible configurations for which no cases are available (known as logical remainders) are used to assist with producing a simplified QCA solution. Software was used to impute outcomes for logical remainders, and this information was accounted for in the QCA solutions, initially generating what was known as a parsimonious solution. The ‘parsimonious solution’ involves the use of an algorithm to impute the likely outcome that would have occurred had the logical remainder been observed. However, in obtaining this solution, some untenable assumptions may have been made in the interest of parsimony, and we generated a further ‘intermediate solution’ that incorporated our own assumptions about the impact of different components (all assumed to be positive in generating a successful outcome). Furthermore, we implemented an algorithm developed by Duşa (2018) to remove untenable and contradictory logical remainders that could be otherwise be used to generate the solution, generating an ‘enhanced intermediate solution’. This solution represented our preferred solution, and is the basis of our interpretation in the results.

### QCA stage 6: Interpreting the solutions

Once we had our QCA solutions we returned to our cases and theory to check that the solutions made sense in the context of individual cases and across cases as a general explanation.

## Results

### Hard mandate studies

QCA revealed that the ‘leading from the front’ theory appeared to explain why some hard-mandate interventions were more successful than others. As the Truth Table (Table 1) below, based on fuzzy-set data, illustrates we had cases for five of the 16 possible configurations. The table illustrates that there is perfect consistency in the relationship between the configuration with all four conditions and cases with the highest levels of vaccine uptake (top row). There is also perfect consistency between higher rates of vaccine uptake and the configuration in which education was absent from the intervention, but the other three conditions were present – although there was only one case with this configuration (second row). The table shows high consistency (0.855) with successful outcomes for the configuration with no two-way engagement but the other three conditions present (row 3, 2 cases). The final two rows illustrate the relationship between configurations associated with unsuccessful outcomes. A configuration in which no intervention components of interest were present, was found in three cases deemed to be mainly unsuccessful and one partially successful case, while a configuration with two components was found in one mainly unsuccessful case. We also emphasise that all the studies achieved statistically significant reductions in the risk of HCWs remaining unvaccinated, and the language of ‘successful’ and ‘unsuccessful’ is relative rather than absolute in this set of results.

Boolean minimisation, and the generation of an enhanced intermediate solution identified two simplified pathways of hard mandate implementation that lead to greater vaccination uptake as illustrated in Table 2 below. The first involves two-way engagement, leadership support and a ‘don’t-go-in-cold’ approach. The second involves leadership support, education and a ‘don’t-go-in-cold’ approach. Therefore, an intervention containing either configuration of components and processes is sufficient to result in a successful outcome. Both configurations cover the majority of instances of the outcome, and crucially they contain all the studies identified as full members of the ‘successful’ outcome set.

**Table 2:** **Minimised intermediate solution for hard mandate QCA**

|  |  | Consistency | PRI | Raw Coverage | Unique Coverage | cases |
| --- | --- | --- | --- | --- | --- | --- |
| **1** | TWOWAYENG*LEADSUP<br>*DONTGOCOLD | 1 | 1 | 0.408 | 0.137 | Stuart;<br>Babcock,<br>Rakita, Smith |
| **2** | LEADSUP *EDUC<br>*DONTGOCOLD | 0.915 | 0.897 | 0.499 | 0.227 | Drees, Frenzel;<br>Babcock,<br>Rakita, Smith |
| **M**<br>**1** |  | 0.932 | 0.921 | 0.636 |  |  |
| TWOWAYENGAGE*LEADSUP *DONTGOCOLD + LEADSUP*EDUC*DONTGOCOLD => SUCCESS |  |  |  |  |  |  |

Notes: See Table 1 for condition names; Upper case conditions indicate the condition is present and lower case indicate a condition is absent; \* = 'AND' relationship; + = 'OR' relationship; Raw coverage: share of outcome covered by a configuration; Unique coverage: share of outcome uniquely covered by a configuration

**Table 3:**
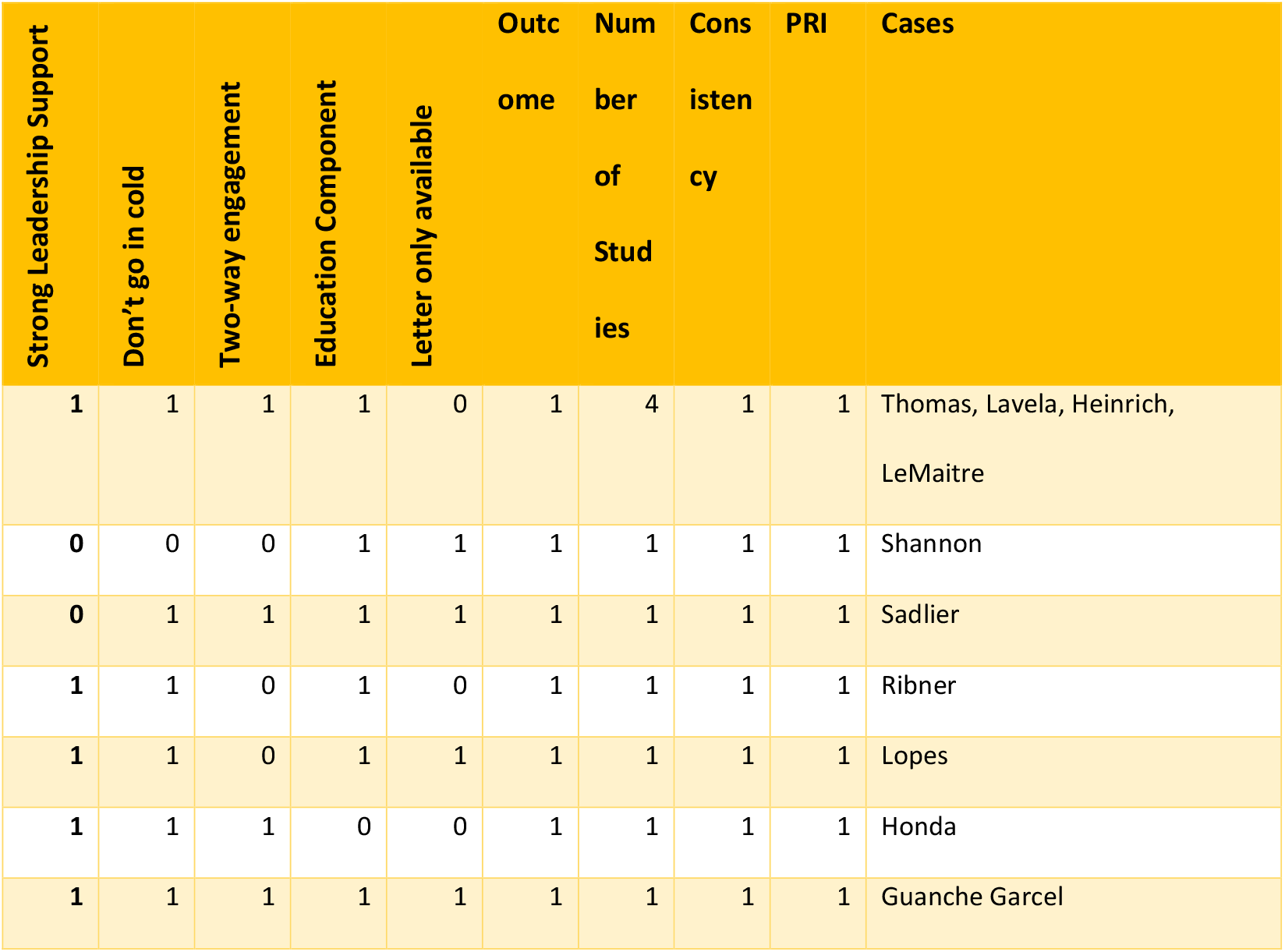
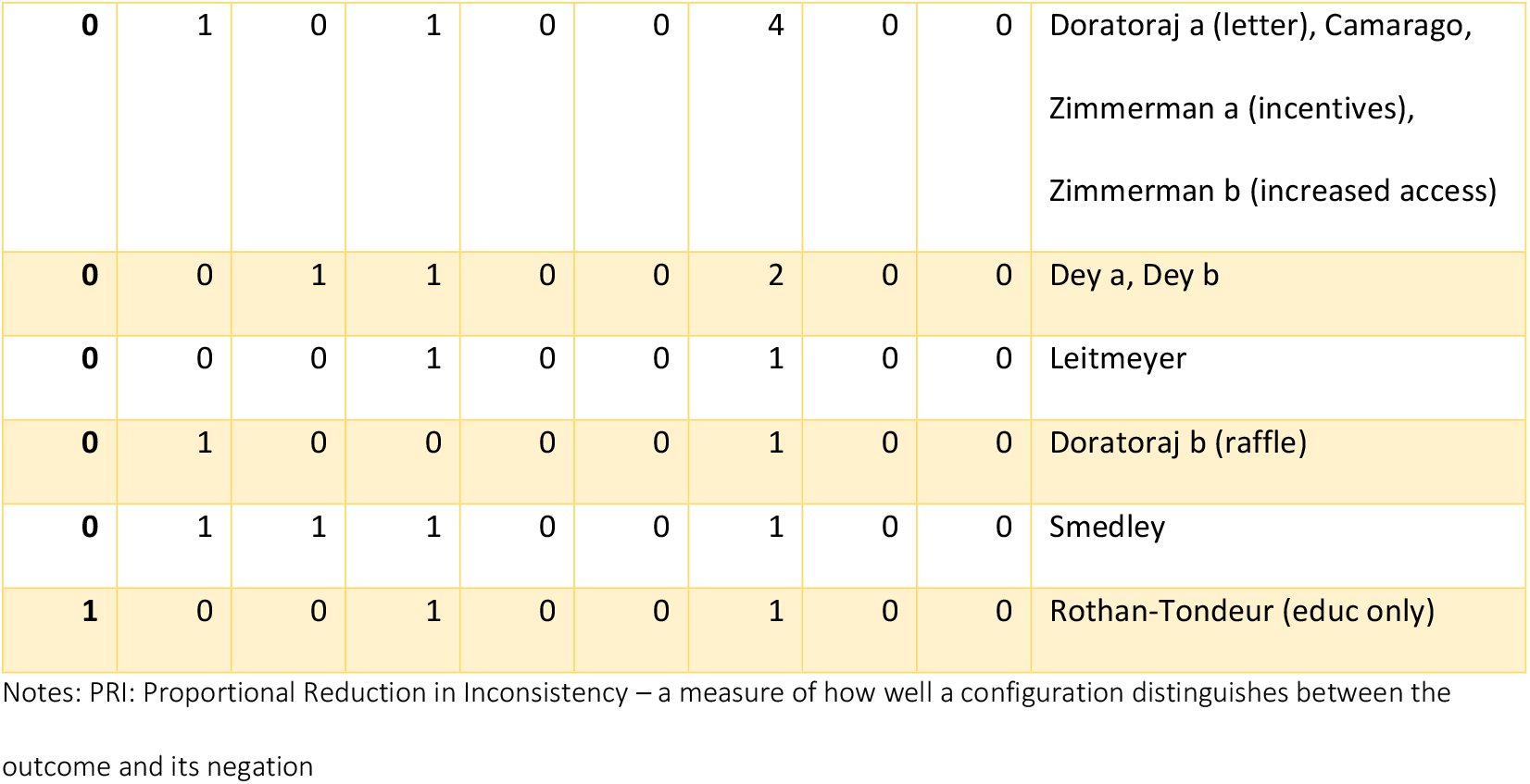
**Truth Table for Soft Mandate / Other QCA (n=20 cases)**

| Strong Leadership Support | Don't go in cold | Two-way engagement | Education Component | Letter only available | Outcome | Number of Studies | Consistency | PRI | Cases |
| --- | --- | --- | --- | --- | --- | --- | --- | --- | --- |
| 1 | 1 | 1 | 1 | 0 | 1 | 4 | 1 | 1 | Thomas, Lavela, Heinrich, LeMaitre |
| 0 | 0 | 0 | 1 | 1 | 1 | 1 | 1 | 1 | Shannon |
| 0 | 1 | 1 | 1 | 1 | 1 | 1 | 1 | 1 | Sadlier |
| 1 | 1 | 0 | 1 | 0 | 1 | 1 | 1 | 1 | Ribner |
| 1 | 1 | 0 | 1 | 1 | 1 | 1 | 1 | 1 | Lopes |
| 1 | 1 | 1 | 0 | 0 | 1 | 1 | 1 | 1 | Honda |
| 1 | 1 | 1 | 1 | 1 | 1 | 1 | 1 | 1 | Guanche Garcel |
| 0 | 1 | 0 | 1 | 0 | 0 | 4 | 0 | 0 | Doratoraj a (letter), Camarago,<br>Zimmerman a (incentives),<br>Zimmerman b (increased access) |
| 0 | 0 | 1 | 1 | 0 | 0 | 2 | 0 | 0 | Dey a, Dey b |
| 0 | 0 | 0 | 1 | 0 | 0 | 1 | 0 | 0 | Leitmeyer |
| 0 | 1 | 0 | 0 | 0 | 0 | 1 | 0 | 0 | Doratoraj b (raffle) |
| 0 | 1 | 1 | 1 | 0 | 0 | 1 | 0 | 0 | Smedley |
| 1 | 0 | 0 | 1 | 0 | 0 | 1 | 0 | 0 | Rothan-Tondeur (educ only) |

Notes: PRI: Proportional Reduction in Inconsistency – a measure of how well a configuration distinguishes between the outcome and its negation

### Soft mandate / other studies (n=20 cases)

The Truth Table below (Table 3) presents configurations using the same four conditions as specified in the ‘leading from the front’ theory, plus an additional condition ‘letter only’. When we first assessed the 20 soft mandate / other cases we had trouble understanding why some highly effective studies did not fit with the theory. It is possible that there are other conditions or contextual factors that explain their success. However, we noticed that these particular studies contained scant information as they were not full research papers but letters only; in particular, they had limited discussion sections which is where critical information, for example about the influence of strong leadership support, was generally reported. Thus, we made the assumption that some of the critical features in the theory were present in these cases but just not described due to the type of article. Once we coded cases as ‘letter only’ (or research articles) and included this in the model, the same patterns began to emerge.

For example, the Truth Table makes clear that all but one of the configurations associated with least effectiveness – in the six bottom rows - did not involve strong leadership support. In contrast, all cases associated with greater effectiveness (aside from two which were letters only) did involve leadership support. Similarly, all cases bar one identified as having a successful outcome had evidence of activities being implemented before the intervention; the one case that did not was a letter.

Boolean minimisation, and the generation of an enhanced intermediate solution identified three simplified pathways of soft mandate and other intervention implementation that led to greater vaccination uptake as illustrated in Table 4 above. These mirror the elements in the solution for hard mandates, with the first two pathways involving conditions around leading from the front and ‘don’t-go-in-cold’. In the first pathway, an additional condition for education was part of the configurations, with the seven studies featuring in this pathway representing a mixture of letters and research articles. In addition to ‘leading from the font’ and ‘don’t-go-in-cold’, the second pathway also includes a condition that is complex to capture within a letter – two way engagement – and unsurprisingly all five cases supporting this pathway were reported in full research articles. The third configuration involved two studies, reported as letters only, with additional conditions representing the absence of reported leadership support and the presence of education. This third pathway consists of two studies where the narrow confines of a letter are unlikely to have allowed for more complex mechanisms and processes such as ‘leaderships support’, two-way engagement, and ‘don’t go in cold’. The data in this QCA model were crisp-set, which facilitated the identification of all instances of the outcome (coverage value of 1) with a coverage score of 1.

**Table 4:** **Minimised intermediate solution for soft mandate / other QCA**

|  |  | Consistency | PRI | Raw Coverage | Unique Coverage | cases |
| --- | --- | --- | --- | --- | --- | --- |
| **1** | LEADSUP*DONTGOCOLD*EDUCATION | 1 | 1 | 0.7 | 0.3 | Ribner; Lopes; Thomas, Lavela, Heinrich, LeMaitre; Guanche Garcel |
| **2** | LEADSUP*DONTGOCOLD*TWOWAYENG*letter | 1 | 1 | 0.5 | 0.1 | Honda; Thomas, Lavela, Heinrich, LeMaitre |
| **3** | leadsup*EDUC*LETTER | 1 | 1 | 0.2 | 0.2 | Shannon; Sadlier |
| **M1** |  | 1 | 1 | 1 |  |  |
| leadsup*EDUC*LETTER + LEADSUP*DONTGOCOLD*EDUCATION + LEADSUP*DONTGOCOLD*TWOWAYENG*letter => SUCCESS |  |  |  |  |  |  |

Notes: See Table 3 for condition names; Upper case conditions indicate the condition is present and lower case indicate a condition is absent; \* = 'AND' relationship; + = 'OR' relationship; Raw coverage: share of outcome covered by a configuration; Unique coverage: share of outcome uniquely coverage by a configuration

Having developed familiarity with the framework and the conditions, we then examined the hard mandates using the crisp-set coding framework developed for the soft mandate/other intervention analysis, and distinguishing those four studies with a RR (<0.2) as (most) successful. Working through the same procedures as the earlier analyses, an enhanced intermediate solution was generated that once again emphasised the importance of ‘leading from the front’, ‘don’t go in cold’ and ‘two-way engagement’ as processes sufficient for generating a successful intervention (Table 5).

**Table 5:** **Minimised intermediate solution for crisp-set hard mandate QCA**

|  |  | Consistency | PRI | Raw Coverage | Unique Coverage | cases |
| --- | --- | --- | --- | --- | --- | --- |
| **1** | TWOWAYENG*LEADSUP*DONTGO<br>COLD | 1 | 1 | 1 | - | Stuart; Babcock,<br>Rakita, Smith |
| **M**<br>**1** |  | 1 | 1 | 1 |  |  |
| **TWOWAYENG\*LEADSUP\*DONTGOCOLD=> SUCCESS** |  |  |  |  |  |  |

Notes: See Table 3 for condition names; Upper case conditions indicate the condition is present and lower case indicate a condition is absent; \* = 'AND' relationship; + = 'OR' relationship; Raw coverage: share of outcome covered by a configuration; Unique coverage: share of outcome uniquely coverage by a configuration

Further checks on the solutions represented in tables 2, 4 and 5 were undertaken. These showed that the solutions did not also trigger the negation of the outcome (a possibility in QCA due to causal asymmetry), and the enhanced intermediate solution generated, using the algorithm developed by Dusa (2018), ensured that untenable simplifying assumptions were not included in deriving our preferred solution.

## Discussion

The above findings reveal that a ‘leading from the front’ rather than a ‘top-down’ approach enhances the effectiveness of flu vaccination drives to increase uptake among HCW. Interestingly, this approach seems to enhance the effectiveness of both hard-mandate approaches and soft-mandates or other approaches. These findings are particularly striking given that the ‘leading from the front logic’ appears to be somewhat in contrast with the overt intervention logic of hard mandates being ‘sticks’ or sanctions to enforce compliance with vaccination drives. By revealing this more nuanced take on hard mandate approaches, our analyses provide additional support for organisations seeking to implement compulsory vaccination drives. Moreover, without this nuanced understanding of key implementation and contextual factors hard mandate approaches may become ineffective in the longer term. And indeed, the lessons learned from these analyses on flu vaccination uptake, may have broader relevance given the twin global concerns of vaccine hesitancy and COVID-19.

Strategies to vaccinate HCWs against infectious diseases have been thrown into sharp relief by the COVID-19 pandemic and the large-scale efforts to vaccinate HCWs against the SARS-CoV-2 virus taking place across countries. Achieving success in campaigns to vaccinate HCWs is of paramount importance as a means of reducing transmission of the virus to vulnerable patients and in order to protect HCWs due to their increased exposure. However, success in vaccinating HCWs is also likely to have broader implications in terms of vaccination uptake, due the influence of HCWs in decisions about vaccination uptake among the general population [48]. The components highlighted here suggest that successful vaccination campaigns among HCWs are dependent on complex conditions, including ‘don’t-go-in-cold’, ‘two-way engagement’ and ‘leading-from-the-front’. Rather than being aligned with any particular model or specific components or activities, these conditions could be considered design principles to be incorporated into future vaccination campaigns. These conditions may also have some salience in considering wider pandemic control measures. In the UK context for example, which at the time of writing has the highest death rate of any large country [49], explanations put forward for non-adherence to pandemic control measures among the general population have parallels with the conditions identified here. For example, the high-profile breach of stay-at-home and social distancing requirements by Dominic Cummins, the Prime Minister’s special advisor, and the subsequent defence of his actions by members of the UK cabinet, has been attributed to weakening adherence to the rules among the population [50]; such actions could be viewed as being in direct opposition to ‘leading-from-the-front’. In contrast, a recent video released by Black UK politicians encouraging vaccine uptake [51], a similar video by British Asian celebrities and politicians [52], as well as the efforts of UK Imams to counter vaccine hesitancy among the UK’s Muslim population [53], can all be viewed as emblematic of ‘leading-from-the-front’.

## Strengths and limitations

This study presents several innovations that help to advance the use of QCA as an evidence synthesis method. First, the QCA drew on a theory developed from the observations of triallists themselves, from the ‘ground up’ and akin to a grounded theory approach. Previous QCA syntheses of systematic review findings have either necessitated drawing on intervention theories derived from logic models with syntheses of process evaluation studies [54], or other separate in-depth qualitative evidence syntheses [16]. The findings here suggest that, in the absence of extant intervention theory or pre-existing synthesis, that working/pragmatic theories can be developed to support QCA synthesis from experiential evidence that is usually overlooked in other synthesis methods, using an ICA framework. Second, this study showed that a theory of how interventions ‘work’, developed through the synthesis of one set of studies using QCA (i.e. the hard mandate studies), can be applied to a conceptually congruent set of separate studies (i.e. the soft mandate and other intervention studies). This form of triangulation can represent a useful adjunct to QCA analyses in systematic reviews that could help to create more robust syntheses in the future. Third, the study also provided a comparison between using fuzzy-set and crisp-set coding schema on the same dataset (hard mandate studies). While similar results were obtained, again providing a further degree of triangulation, the fuzzy-set coding for the hard mandate studies was a more appropriate choice conceptually. This was with respect to both the coding for the outcome, where all the studies had obtained significant reductions in unvaccinated (despite heterogeneity in the original meta-analysis [9]), as well as the conditions, where in the case of ‘don’t go in cold’ in particular, different levels of previous engagement were apparent among some hard mandate studies in a way which wasn’t as apparent for studies on soft mandates and other intervention modes. Fourth, this is the first example that we are aware of where ‘publication type’ was included in the analysis and was predictive of outcomes. This work thus provides some evidence in support of one issue that’s been long suspected in systematic reviews: that the lack of information in some papers / publications can lead to unreliable review results – and possibly undermine other subgroup analyses [55]. Finally, this study once again is further demonstration of the potential for further adjunct analysis of evidence that has already been assembled and synthesised in some form, to address new questions and generate new understandings. This study drew on ICA/QCA; other techniques for the reanalysis of existing review evidence have also been suggested elsewhere [56]. Given the large volume of systematic reviews being published annually, each requiring substantial investment and sometimes generating conflicting results or interpretations, techniques for further probing of the included studies to provide additional nuance or address questions not considered by the original reviewers, may continue to develop as a promising adjunct stream of evidence synthesis.

While the analyses presented here are of importance, both in (i) revealing some of the conditions sufficient to result in successful influenza vaccination campaigns: as well as (ii) emphasising the potential of ICA/QCA in enhancing our understanding of existing review evidence, some limitations should be noted. An important limitation is around the approach itself and its capacity to consistently and correctly reveal complex causal relationships. There exist some critiques around the potential of QCA to produce correct solutions in simulation studies [57], although responses provided by others not only highlight flaws in these critiques, but also emphasise that a QCA solution cannot be generated and articulated in the absence of case and substantive knowledge [58]. While we regard the use of ICA to generate theory to underpin QCA as a useful innovation in the field; we nevertheless recognise that trial reports remain sparse in terms of reporting intervention details [12], and despite the allowances we made for sparse reporting in letters, ‘missing data’ may be a further caveat on the results. Finally, while we generated an enhanced intermediate solution, following procedures developed by Duşa [47], the treatment of logical remainders somewhat contested and unresolved in the literature [59], which could represent a final caveat to these results. However, since QCA requires that the solution is consistent with a programme theory that is identifiable in all relevant cases, it can be seen, in some ways, as having a higher bar for achieving a credible explanation than statistical analysis. In a statistical analysis, deviant cases might increase variance / confidence intervals, but are considered ‘explained’ when this happens. In a QCA, a deviant case indicates that a credible solution that properly explains what is going on has not be found, so further analysis is required. As such, given that we identified consistent patterns of association across several independent research studies and that the detail of each case was consistent with our ‘leading from the front’ theory, the credibility of these findings is strengthened.

## Conclusion

Regardless of intervention type a ‘leading from the front’ approach to implementation, which incorporates building on institutional knowledge, education, opportunities for two-way engagement and strong leadership support, will likely enhance the success of HCW flu vaccination drives. While the results pertain to flu vaccination and HCW populations, the nuanced understanding of effective intervention strategies identified here may be useful in the urgent efforts to vaccinate HCW and the general public against COVID-19.

## Data Availability

All data is made available within the article and supplementary file.

## Acknowledgements

This paper was initially developed as part of a small project funded by the UCL Global Engagement Fund on ‘Improving our understanding of how interventions ‘work’ in Oceania’ and builds on reviews and methodological work conducted for The London/York NIHR Policy Research Programme Reviews Facility. We are very grateful for all the support we received from Associate Professor Cath Chamberlain at Latrobe University, who made the project possible, as well as the input from participants at QCA workshops held in Melbourne (Latrobe) and Adelaide. Finally, this study represents a re-analysis of previous work by Theodore Lytras and colleagues (2016) and Theo Lorenc and colleagues (2017). This study could not have been conducted without the diligent work of both author teams which preceded this study and facilitated re-analysis of this literature. However, neither the Lyras or Lorenc team were involved in the study reported here, and the views and results expressed are those of the authors and not necessarily those of the Lytras or Lorenc author teams, the NHS, the NIHR, the Department of Health and Social Care, or its partners.

## Supporting information

### Hard mandate (Analysis 1) Fuzzy set Data table

| Studies | Outcome set | Don't go in cold (DONTGOCOLD) | Educational Component (EDUC) | Information other (INFORM)* | Two-way engagement (TWOWAYENG) | Stigma (STIGMA)* | Strong Leadership Support (LEADSUP) | Multi-setting (MULTI)* |
| --- | --- | --- | --- | --- | --- | --- | --- | --- |
| Awali | 0.33 | 0.33 | 0 | 0.66 | 0 | 0 | 0 | 0 |
| Babcock | 1 | 1 | 1 | 0 | 0.66 | 0 | 1 | 0.66 |
| Drees | 0.66 | 1 | 1 | 0.66 | 0 | 1 | 0.66 | 0.33 |
| Frenzel | 0.66 | 1 | 1 | 0.66 | 0 | 0.66 | 1 | 0.33 |
| Hospital-Ksienski | 0.33 | 0 | 0 | 1 | 0 | 0.66 | 0 | 1 |
| ResiCare-Ksienski | 0.33 | 0 | 0 | 1 | 0 | 0.66 | 0 | 1 |
| Leibu | 0.66 | 0 | 0 | 0.33 | 0 | 0 | 0 | 0.66 |
| Podscervinski | 0.33 | 1 | 0 | 0.66 | 0 | 0.33 | 1 | 0 |
| Rakita | 1 | 0.66 | 1 | 0.66 | 1 | 0 | 1 | 0.66 |
| smith | 1 | 0.66 | 1 | 0.66 | 0.66 | 0 | 1 | 0.66 |
| stuart | 1 | 1 | 0 | 0.33 | 1 | 0 | 1 | 0.33 |

\*Not included in final models

### Soft mandate/other (Analysis 2) Data table

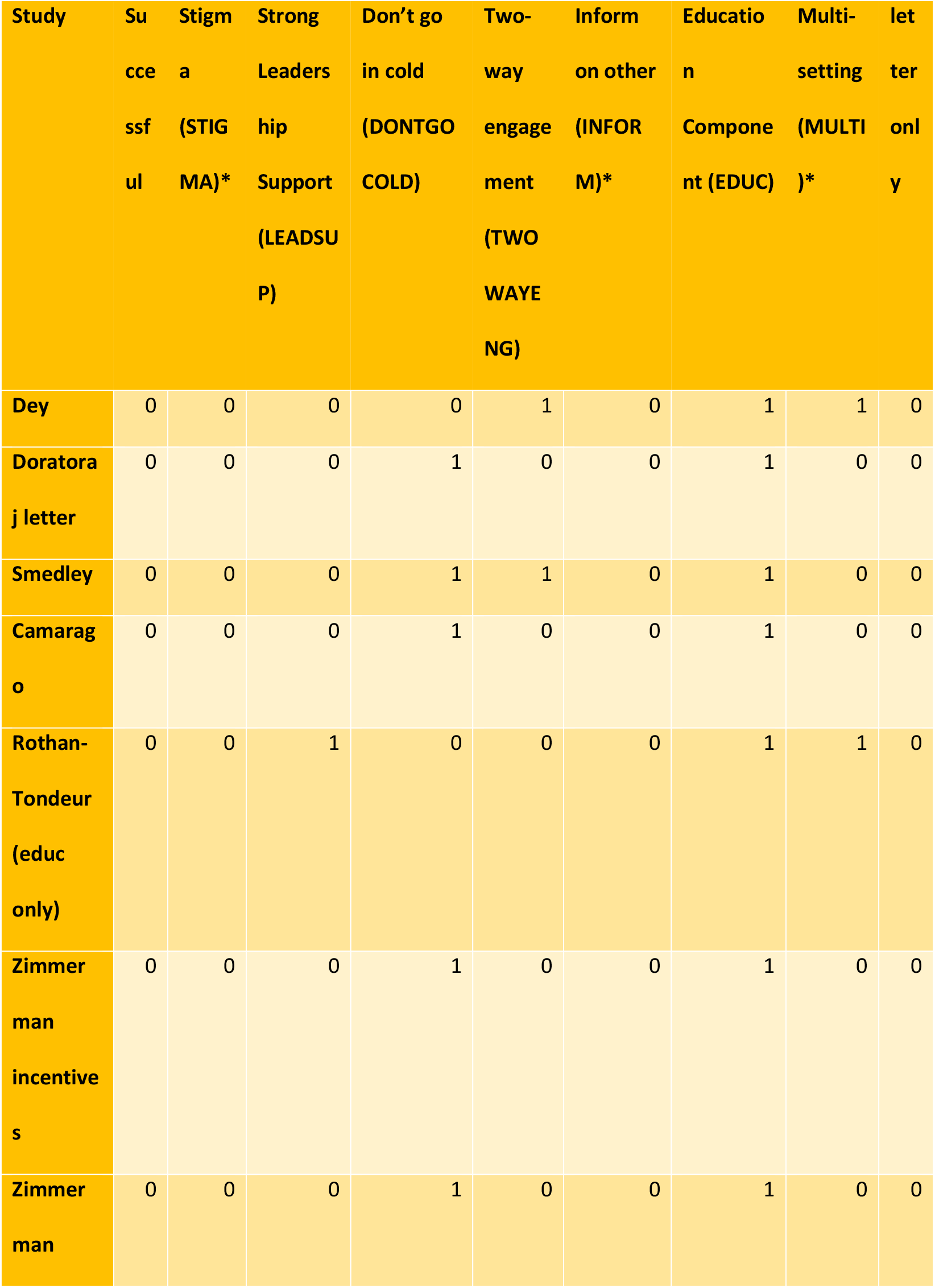
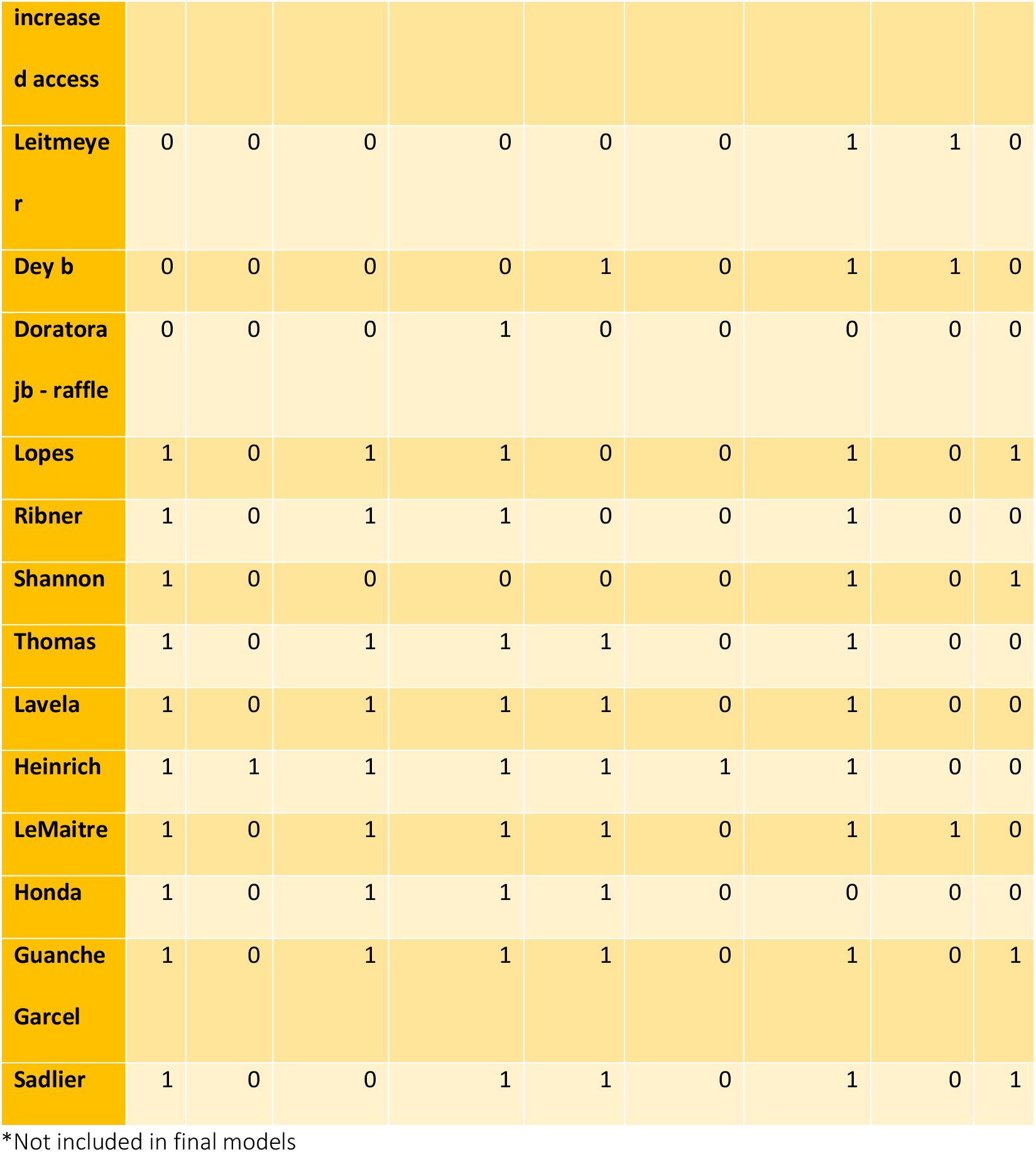

### Hard mandate (Analysis 1) Crisp set Data table

| Studies | RR | Outcom<br>e set | Don't go in<br>cold<br>(DONTGOCOLD<br>) | Education<br>Componen<br>t (EDUC) | Two-way<br>engagement<br>(TWOWAYENG<br>) | Strong<br>Leadershi<br>p Support<br>(LEADSUP<br>) |
| --- | --- | --- | --- | --- | --- | --- |
| Awali | 0.35 | 0 | 1 | 0 | 0 | 0 |
| Babcock | 0.06 | 1 | 1 | 1 | 1 | 1 |
| Drees | 0.24 | 0 | 1 | 1 | 0 | 1 |
| Frenzel | 0.21 | 0 | 1 | 1 | 0 | 1 |
| Hospital-Ksienski | 0.44 | 0 | 0 | 0 | 0 | 0 |
| ResiCare-Ksienski | 0.57 | 0 | 0 | 0 | 0 | 0 |
| Leibu | 0.22 | 0 | 0 | 0 | 0 | 0 |
| podscervinsci | 0.42 | 0 | 1 | 0 | 0 | 1 |
| Rakita | 0.05 | 1 | 1 | 1 | 1 | 1 |
| smith | 0.08 | 1 | 1 | 1 | 1 | 1 |
| stuart | 0.14 | 1 | 1 | 0 | 1 | 1 |

### Hard Mandate (Analysis 1) Crisp Set Truth table

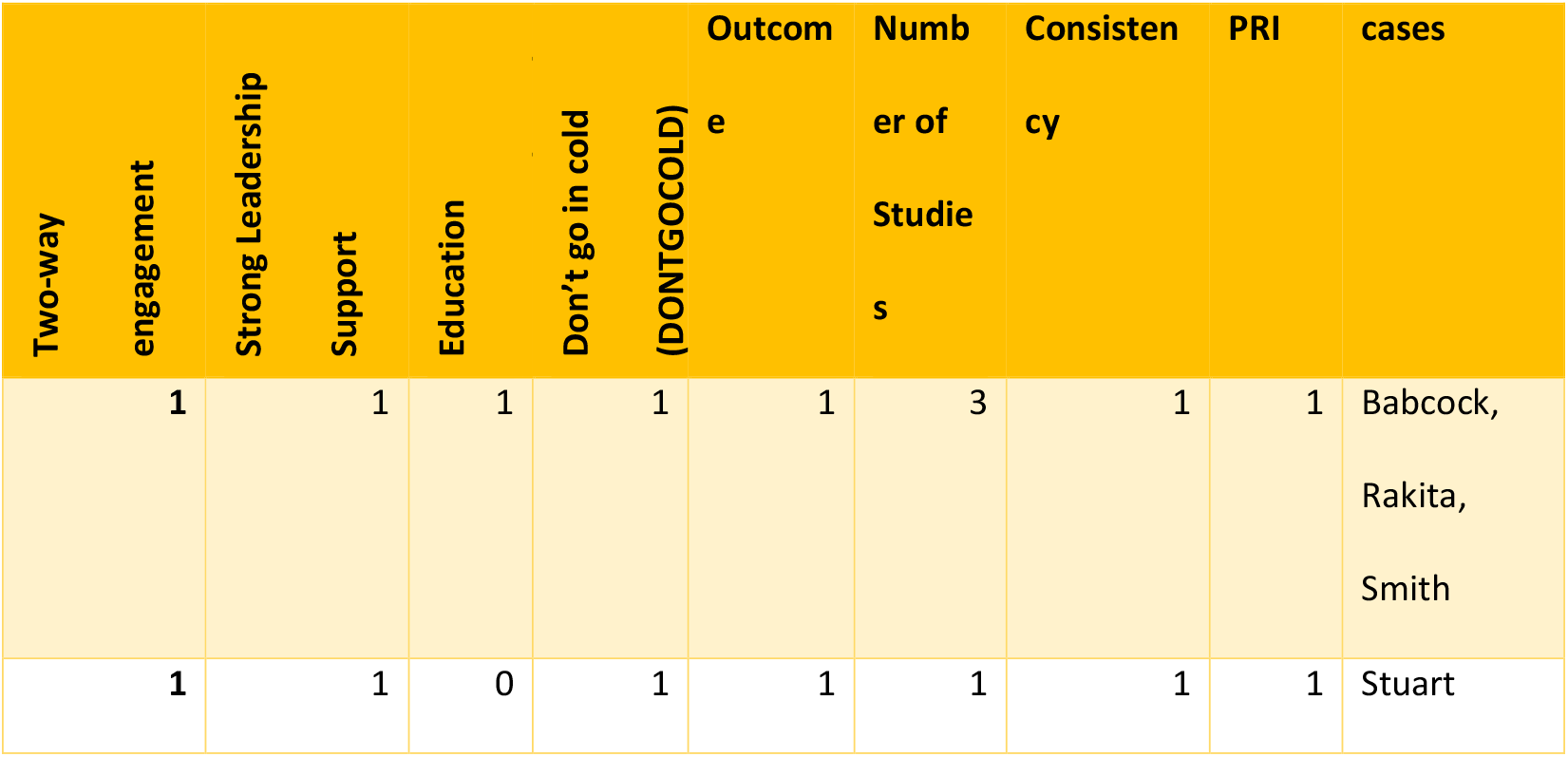
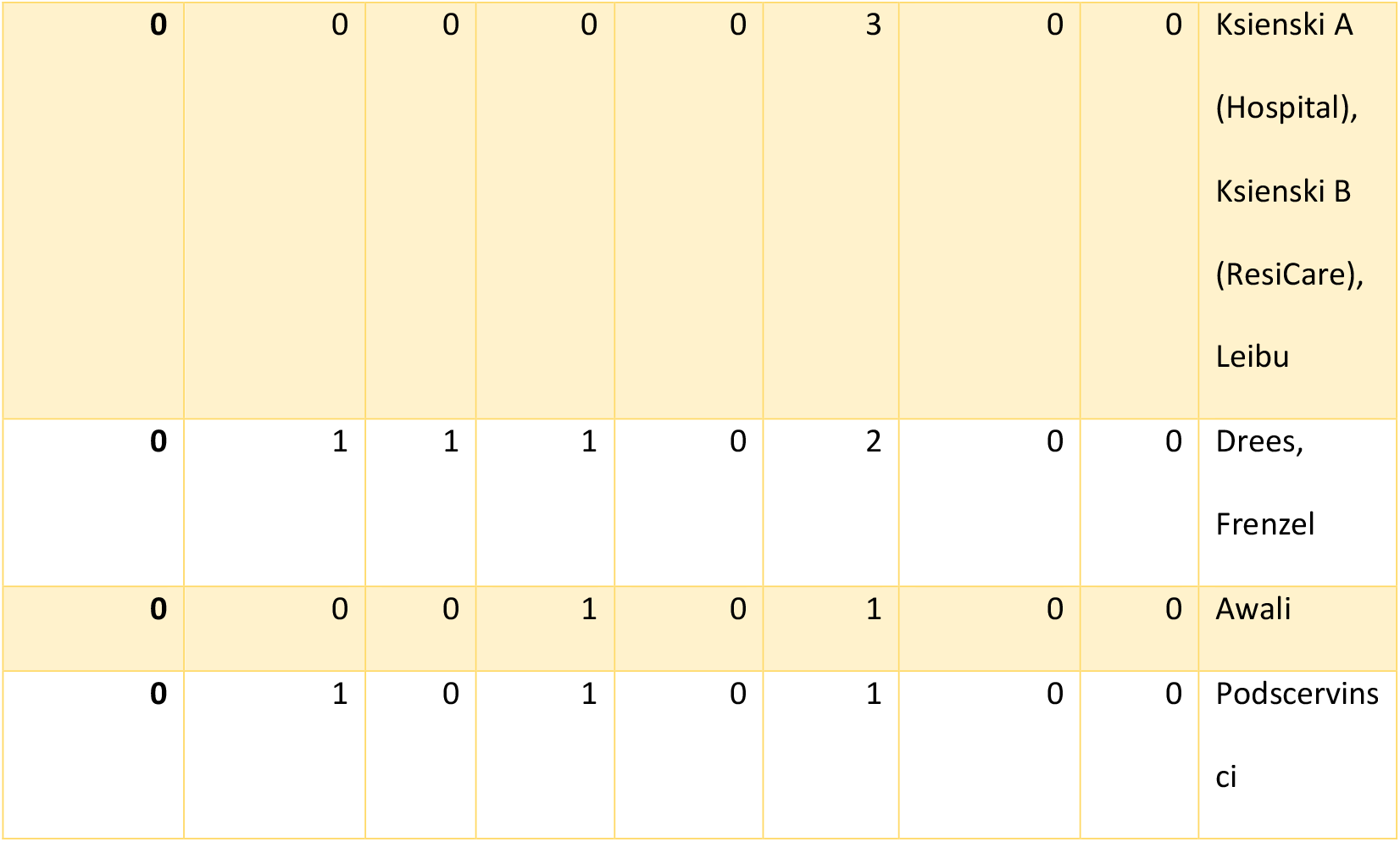

### Hard mandate data table with evidence

|  |  |  |  | 'Don't go in cold'<br>(Have there been other previous vaccination campaigns prior to current hard mandate intervention to change behaviours) | Was education provided to improve knowledge among HCW of vaccination benefits and risks? | Were Healthcare Workers encouraged to inform on others? | Two-way engagement/bidirectional communication | Stigma | Area policies implemented across institutions | Leading from the front – did senior staff engage in the campaign |
| --- | --- | --- | --- | --- | --- | --- | --- | --- | --- | --- |
| Studies | Risk Ratio | Percentage of HCWs vaccinated at the end of intervention | Outcome set value | 0 – No effort mentioned<br>0.33 – Sanction-based or accountability-based efforts only previously<br>0.66 – Incentivisation and health promotion | 0 – No education provided or education did not improve levels of knowledge<br>1 – Education provided that improved knowledge of the vaccine | 1 – Employees encouraged to inform on colleagues<br>0.66 – Managers observed compliance<br>0 – No evidence of policy implemented across all employees | 0 - not stated<br>0.66 - engagement not continuously sought and no formal system - ad hoc<br>1 - formal processes for two way engagement established | 0 – not stated<br>0.33 – stigmatising process of declination<br>0.66 – public display of vaccination status enforced only<br>1 – stigmatising language with | 0 – single institution<br>0.33 – multiple institutions and/or number of employees >1,500<br>0.66 – multicentre institutions<br>1 – Area based interventions (e.g. states or counties) | 0 – not stated<br>0.66 – leadership approved and facilitated campaign<br>1 – leadership engaged in and publically voice support for vaccination campaign OR described as being |

|  |  |  |  | based efforts<br>previously<br>1 – Combination of<br>sanction and<br>incentive-based<br>measures used prior<br>to current<br>intervention |  |  |  | public display of<br>vaccination status |  | instrumental for<br>success |
| --- | --- | --- | --- | --- | --- | --- | --- | --- | --- | --- |
| Awali | 0.35 | 93 | 0.33 | Value: 0.33;<br>Previous efforts<br>were sanction based<br>only “During the<br>next season (2010-2011), the<br>institutional<br>vaccination policy<br>was not strictly<br>mandatory;<br>however, all<br>unvaccinated<br>employees were<br>required to wear masks when within 6 feet of all patient contacts.” | Value: 0; “The<br>reluctance of some<br>HCP at our hospital<br>to receive the<br>influenza vaccine<br>despite the<br>mandatory<br>vaccination policy<br>most likely reflects<br>misperceptions and<br>poor knowledge of<br>the benefits and risks of the vaccine.<br>“ | Value: 0.66;<br>Managers monitored<br>compliance “The<br>OHS and HCP’s<br>direct supervisors or<br>managers are<br>responsible for<br>ensuring compliance<br>with this policy.” | Value 0; No evidence | Value 0; No<br>evidence of<br>stigmatising<br>processes | Value 0; “...a cross-<br>sectional survey<br>research study was<br>conducted at an<br>urban tertiary care<br>hospital in the<br>metropolitan Detroit<br>area” | Value 0; not stated |
| Babcock | 0.06 | 98.4 | 1 | Value: 1; Non-sanction based campaigns initially before accountability-based measures imposed: “free vaccine available at multiple sites and times, extensive publicity, incentives and educational programs, and more recently, declination statements..... In 2007, influenza vaccination rates were added to the BJC patient safety and quality scorecard used at all hospitals in the organization..... In 2008, BJC HealthCare implemented a mandatory influenza vaccination policy for all employees” | Value: 1; “Key factors that supported the success of the program included consistent communication emphasizing patient safety and quality of care, coordinated campaigns, leadership support, and medical director support to talk with any employee with concerns about the vaccine, on request.” | Value: 0; No specific enforcement or data on compliance collected | Value 0.66; Ad hoc interactions: “Managers interacted with their staff to ascertain reasons for noncompliance and to provide coaching about influenza” | Value 0; No evidence of stigmatising processes; encouraged to wear mask | Value 0.66; “Facilities include 11 acute care hospitals and 3 extended care facilities, as well as day care centers, employed physician groups, occupational medicine, home care, and behavioral health services.” | Value 1; “The CEO of BJC published a letter in the BJC newspaper explaining the rationale for the policy. The multidisciplinary implementation team met regularly before and during the vaccination campaign to ensure timely, consistent, and coordinated communication and responses to any issues that arose.” |
| **Drees** | **0.24** | **92** | **0.66** | **Value: 1;** Non-sanction based campaigns initially escalating to other forms of mandates/sanctions : “[the] vaccination campaign included promotional materials, web- | **Value: 1;** from supplementary materials, the communication campaign included: “Launched internal flu website with explanation of new program, frequently asked questions, | **Value: 0.66;** Managers monitored compliance “Beginning 2 weeks after the start of the campaign, every manager and vice president in the system began receiving weekly | **Value 0;** No evidence | **Value 1;** After vaccination (or attesting vaccination elsewhere), HCP were given hanging badges, stating “I’m vaccinated because I care,” to wear with their regular identification | **Value 0.33;** “Christiana Care Health System is a 2-hospital, 1,100-bed, private, not-for-profit, community-based academic healthcare system located in northern Delaware.” | **Value 0.66;** At each entrance, volunteer “clerks” (who ranged from administrative assistants to leadership personnel) scanned the HCP’s identification badge |
|  |  |  |  | based and in-person education, free vaccination for employees and medical-dental staff, roving vaccinators...During the 2009 H1N1 pandemic, the health system created a policy that required explicit declination by all employees as well as the wearing of surgical masks....However, the policy did not include provisions to enforce either of these measures.” | multiple resources and links to external sources; Web-based education (non-mandatory) to all employees” | lists of their employees, notated as vaccinated, not vaccinated, or no response.” |  | badges. Wearing the tag was not mandatory, but anyone not wearing an “I’m vaccinated” tag was required to mask while in patient care areas, regardless of their actual vaccination status. |  | and the appropriate form (taking ~30 seconds), and then directed him/her to the next available vaccinator (volunteer nurses and pharmacists). ... Health system leadership approved use of the employee influenza vaccination rate as 1 of 3 metrics comprising a pre-existing employee bonus program, known as the Transformation |
|  |  |  |  |  |  |  |  |  |  | Rewards Program (TRP). |
| **Frenzel** | **0.21** | **94** | **0.66** | Value: 1; Non-sanction based campaigns initially before mandatory: “the employee influenza vaccination program consisted of large, on-site influenza vaccination clinics that were distributed throughout >20 geographically dispersed patient care areas and research and administration buildings and were supplemented by 1 | Value: 1; “We expanded our education and communication campaign by prominently advertising the expanded clinic schedule and centralized, hospital-based locations and distributing various educational materials on the safety and efficacy of influenza vaccination.” | Value: 0.66; Managers monitored compliance “Compliance with mask use for unvaccinated HCWs was the responsibility of supervisors in each clinical area and was documented in a vaccine preventable diseases policy compliance-monitoring database” | Value 0; No evidence | Value 0.66; “compliance stickers also promoted positive reinforcement from co-workers and patients who perceived vaccination as an important patient safety measure.” | Value 0.33; “The University of Texas MD Anderson Cancer Center is a 656-bed National Cancer Institute–designated comprehensive cancer center with >19,000 employees.” | Value 1; “Senior leadership supported our initiative by aligning institutional goals with the 2007 Joint Commission requirement to increase HCW influenza vaccination rates.” |
|  |  |  |  | week of roaming vaccination services via mobile carts... in 2009, we piloted the mandatory participation influenza prevention program, which targeted HCWs in high-risk areas and in the nursing staff as subsequently defined” |  |  |  |  |  |  |
| **Ksienski**<br>**(a)** | **0.44** | **74** | **0.33** | **Value: 0;** No evidence of activity before hard mandate imposed | **Value 0;** No evidence of substantial education measures alongside punitive measures | **Value: 1;** Employees encouraged to inform on one another “HCWs who witness any colleagues violating the Policy are required to report<br>the incident to their supervisor” | **Value 0;** No evidence | **Value 0.66;** Green dot stickers used to publically indicate vaccination status | **Value 1;** “province-wide Influenza Prevention Policy, whose primary objective is to increase vaccination coverage rates of HCWs.” | **Value 0;** not stated |
| **Ksienski**<br>**(b)** | **0.57** | **75** | **0.33** | **Value: 0;** No evidence of activity before hard mandate imposed | **Value 0;** No evidence of substantial education measures alongside punitive measures | **Value: 1;** Employees encouraged to inform on one another “HCWs who witness any colleagues violating the Policy are required to report the incident to their supervisor” | **Value 0;** No evidence | **Value 0.66;** Green dot stickers used to publically indicate vaccination status | **Value 1;** “province-wide Influenza Prevention Policy, whose primary objective is to increase vaccination coverage rates of HCWs.” | **Value 0;** not stated |
| **Leibu**<br>**and**<br>**Maslow** | **0.22** | **94.7** | **0.66** | **Value: 0;** No evidence of activity before hard mandate imposed | **Value 0;** No evidence of substantial educational activities that could address employee concerns | **Value: 0;** No evidence | **Value 0;** No evidence | **Value 0;** Nothing mentioned | **Value 0.66;** “AHS comprised three acute care adult hospitals, a children’s hospital, an inpatient rehabilitation hospital, home care, transportation services, and several off-site clinical office practices including diagnostic facilities.” | **Value 0;** not stated |
| **Podscer**<br>**vinsci** | **0.42** | **96** | **0.33** | **Value: 1;** Non-sanction based campaigns initially before mandatory measures (note different intervention strategies had been implemented – evidence of one provided): “vaccine availability was advertised via multiple modalities at the center, including; mass emails, newsletter articles, and intranet postings. All employees were required to either be vaccinated or to complete a one-page signed declination form acknowledging that they understood the risks of declining the vaccine in a setting with such high-risk patients” | **Value 0;** Education only followed after declination as a penalty: “required decliners to complete enhanced influenza vaccine education” | **Value: 0.66;** Managers monitored compliance “Staff that did not meet campaign deadlines by either receiving or declining the vaccination were required to meet with their respective manager” | **Value 0;** No evidence | **Value 0.33;** In-person declination process “in front of occupation health, infection prevention staff” | **Value 0;** “The study was performed at a large comprehensive cancer care center...” | **Value 1;** “Center leadership support/involvement” described |
| **Rakita** | **0.05** | **98.9** | **1** | **Value: 0.66;** Non-sanction based campaigns initially before mandatory measures (note different intervention strategies had been implemented – | **Value 1;** “In the spring of 2005, multiple focus groups of staff and managers were created to gather data on the barriers, educational deficits, and preferences in receiving | **Value: 0;** No evidence as a policy across all employees | **Value 0;** No evidence | **Value 0;** Nothing mentioned | **Value 0.66;** “a tertiary care, multispecialty medical center that includes a 336-bed hospital, adjoining outpatient clinics, 7 regional clinics, and a research center, that provides | **Value 1;** Intervention included “meetings with staff and leadership to answer questions; grand rounds speakers; trained advocates, or “champions,” of |
|  |  |  |  | evidence of one provided): “vaccine | information with regard to vaccinations. The campaign was organized around the information gathered during these focus-group sessions.” |  |  |  | residency teaching programs, and that employs approximately 400 physicians and a total of approximately 5,000 HCWs.” | influenza vaccination; and one-on-one meetings with concerned staff. These champions included the president and CEO of the medical center.” |
| **Smith** | **0.08** | **97.7** | **1** | **Value: 0.66;** Non-sanction based campaigns initially before mandatory measures: During 2009 pandemic, prior to 2011 mandates, the hospital “engaged in unprecedented community and internal publicity, | **Value 1;** “engaged in unprecedented community and internal publicity, education, and other efforts to improve HCW influenza vaccination rates” | **Value: 0.66;** Managers monitored compliance “Managers were responsible for monitoring the vaccination/exempti on status of employees in their department” | **Value 0;** No structured process for encouraging interactions invited: “Aurora has a formal process for measuring HCW job satisfaction, but the process did not include questions about the vaccination policy.” | **Value 0;** Nothing mentioned | **Value 0.66;** “Aurora Health Care (Aurora) is a large integrated delivery system in eastern Wisconsin/northern Illinois that serves over 1.2 million patients per year and has over 30,000 employees....BJC Healthcare, a large | **Value 1** “Senior leadership support was critical to the program’s success and its continuation.” |
|  |  |  |  | education, and other efforts to improve HCW influenza vaccination rates” |  |  |  |  | Midwestern health care organization similar in size and revenue to Aurora” |  |
| **Stuart** | **0.14** | **92.8** | **1** | **Value: 1;** Incentive-based programme implemented before a sanction-based programme was trialled before hard mandate intervention: “The program is free and incorporates mobile rounds, extended hours and promotion via newsletters and announcements. .... In December 2012, the DN was<br>informed that to increase influenza vaccination rates, unvaccinated HCWs would be asked to wear a surgical mask during patient care throughout the influenza season.<br>Staff were given the opportunity to ask questions about the program and raise any concerns. In February 2013, a follow-up letter confirmed that the program would be enforced, and vaccination commenced in April 2013 (when the<br>vaccine became available). | **Value 0;** “No evidence of substantial educational activities” | **Value: 0;** No evidence | **Value 1;** “Staff were given the opportunity to ask questions about the program and raise any concerns.” | **Value 0;** Nothing mentioned | **Value 0.33;** “Monash Health is a tertiary referral service in Melbourne, Australia, with 2200 beds and 13 389 HCWs. The service provides for 1.3 million residents.” Note intervention described as being carried out in one department; unclear how many HCWs involved. | **Value 1;** Senior leaders on authorship team |

### Soft mandate/other data table with evidence

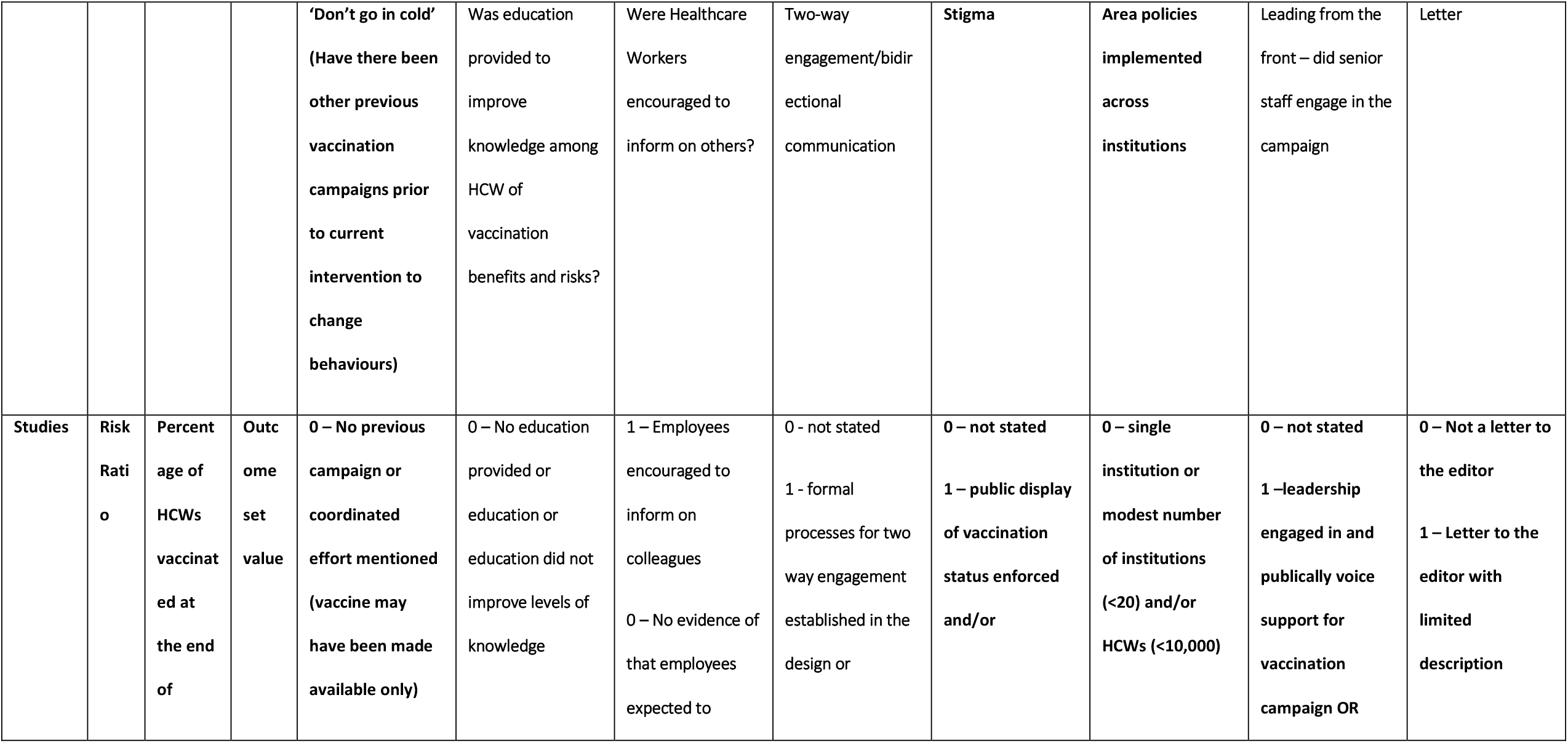
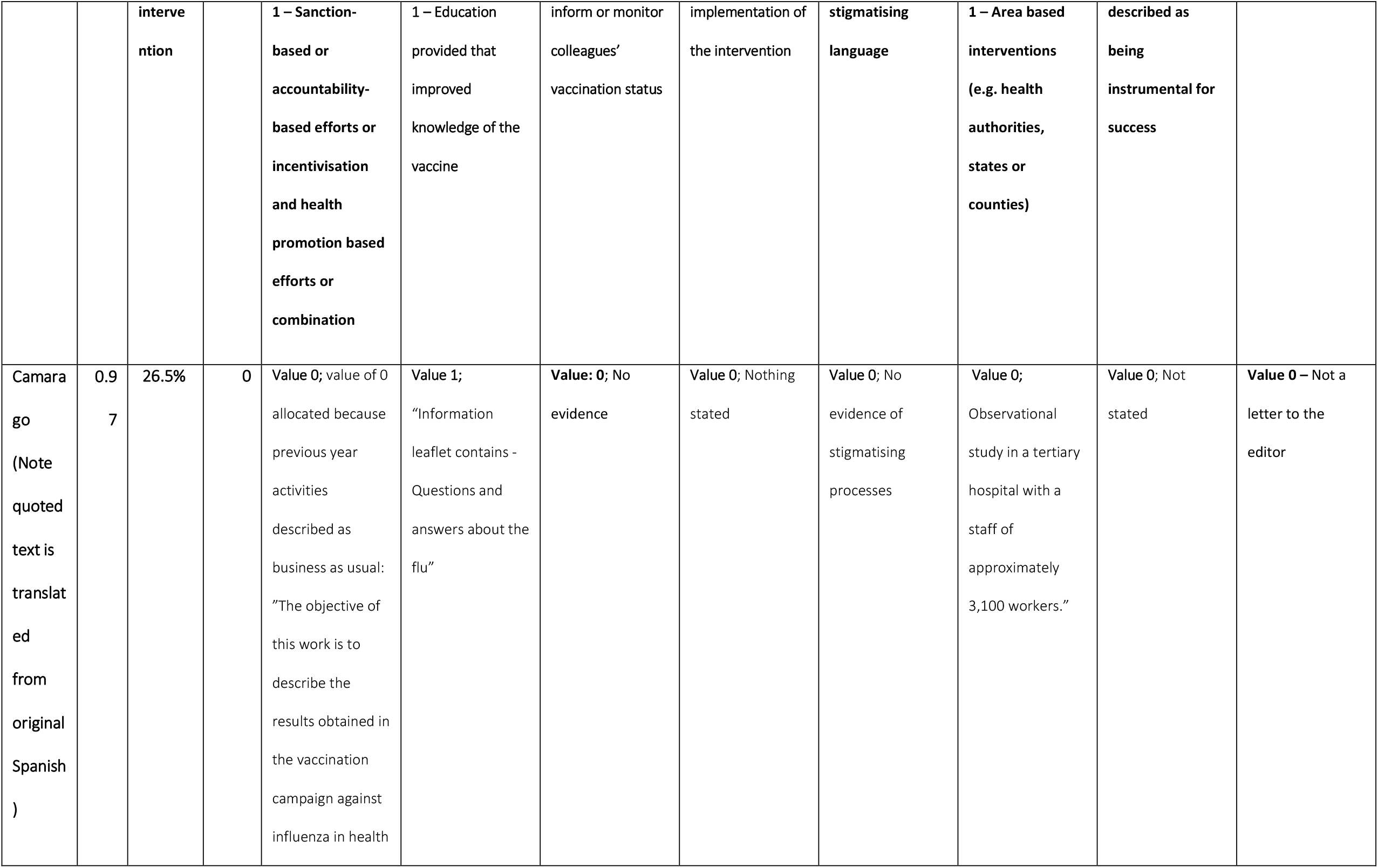
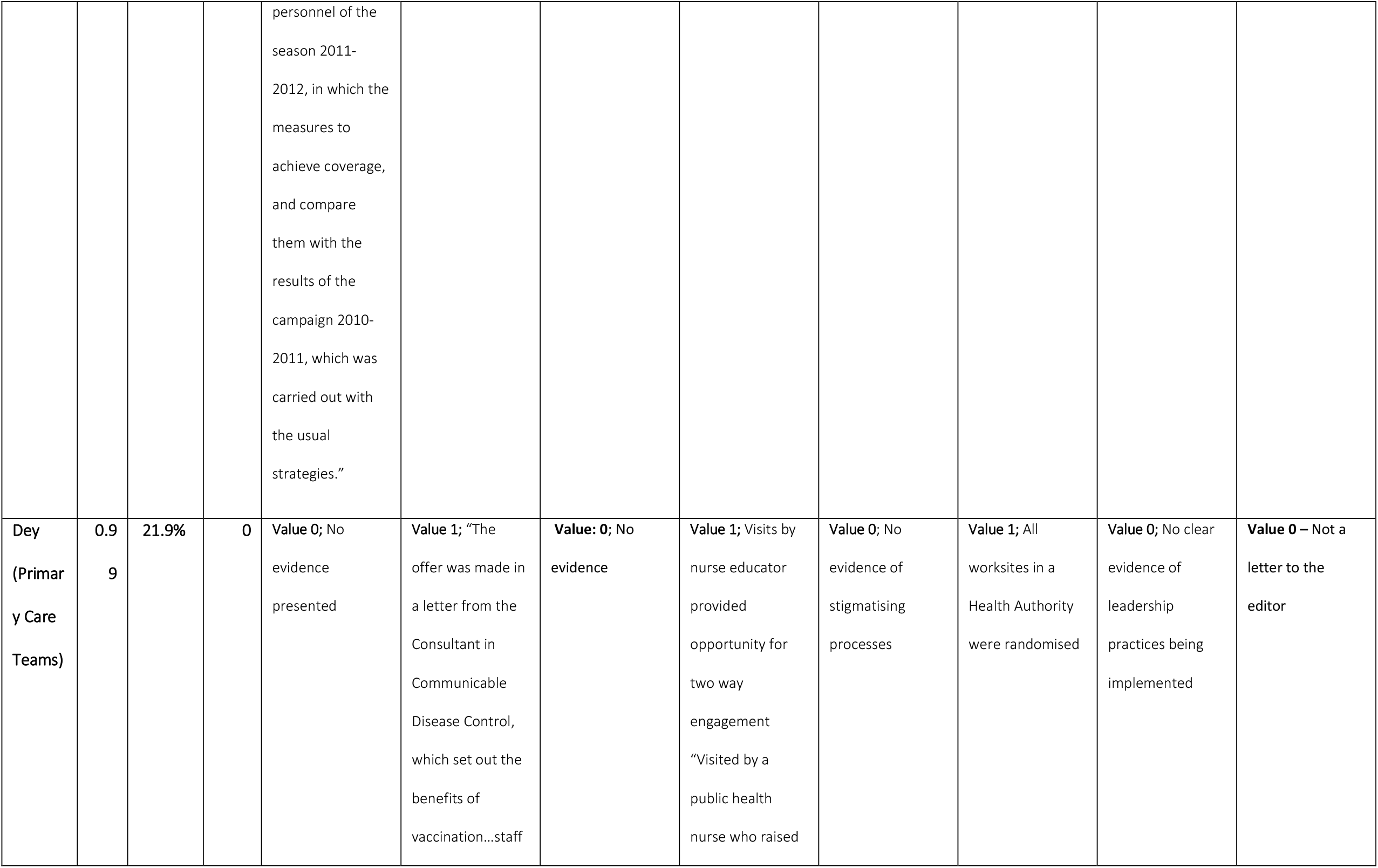
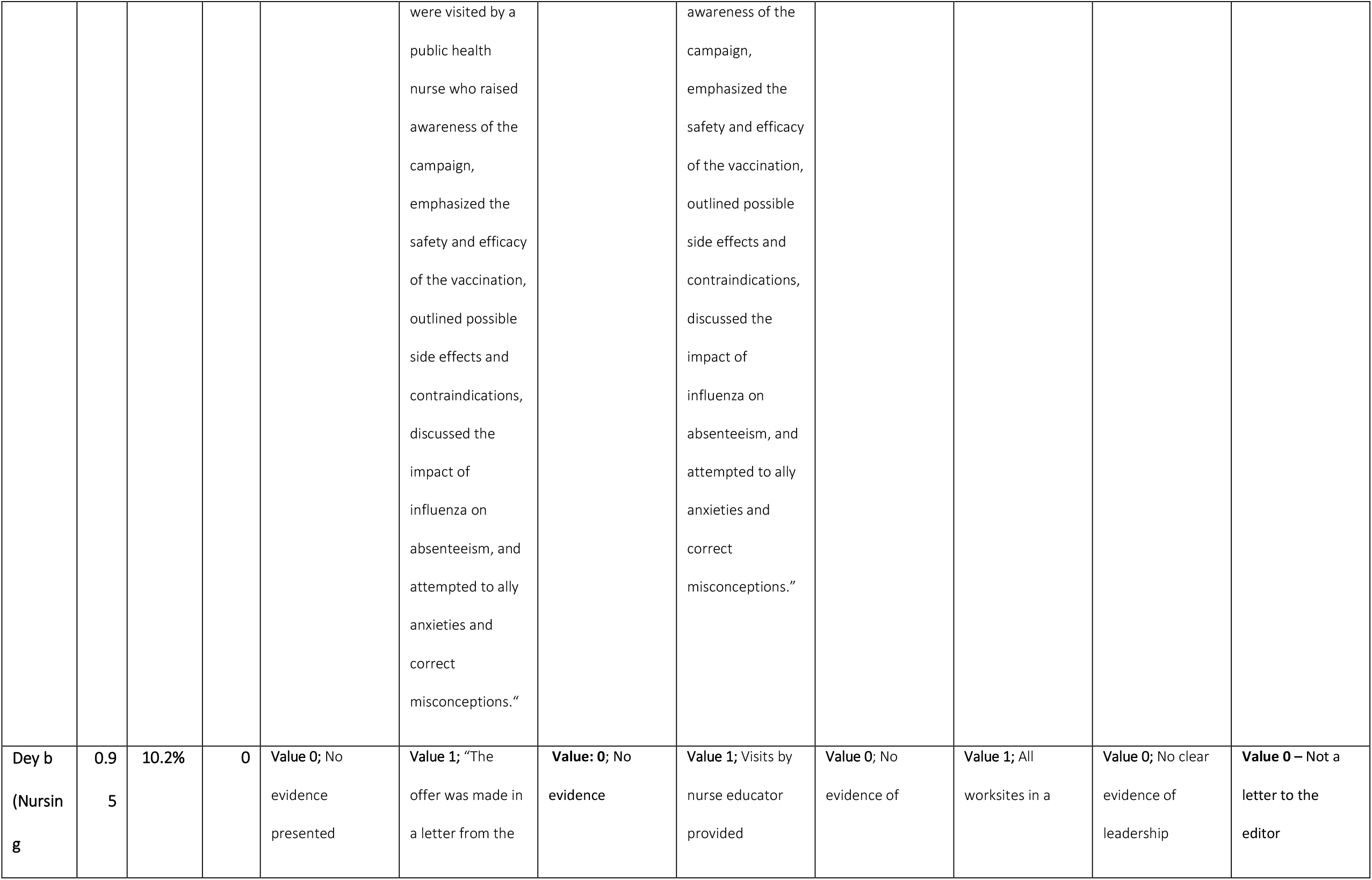
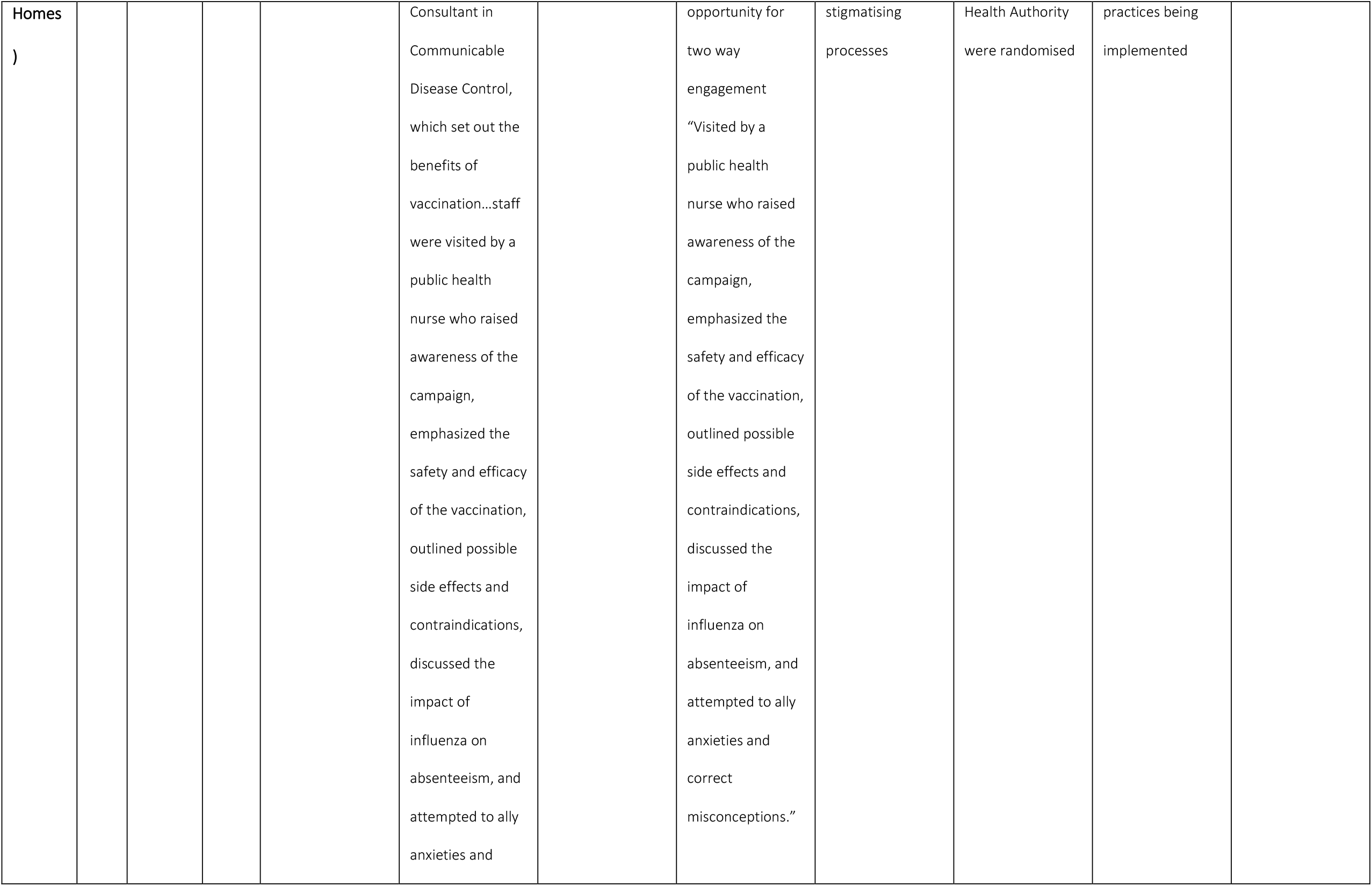
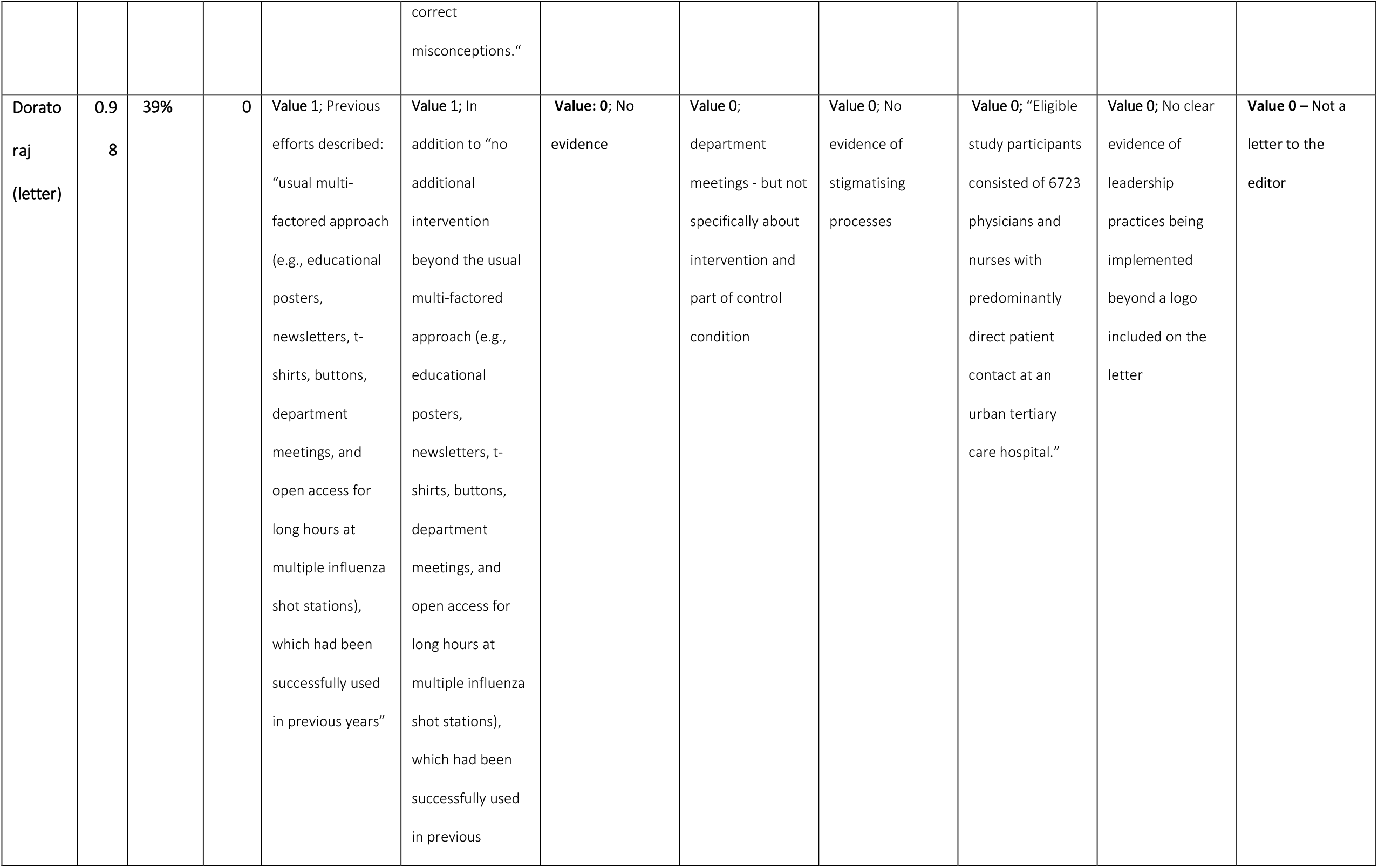
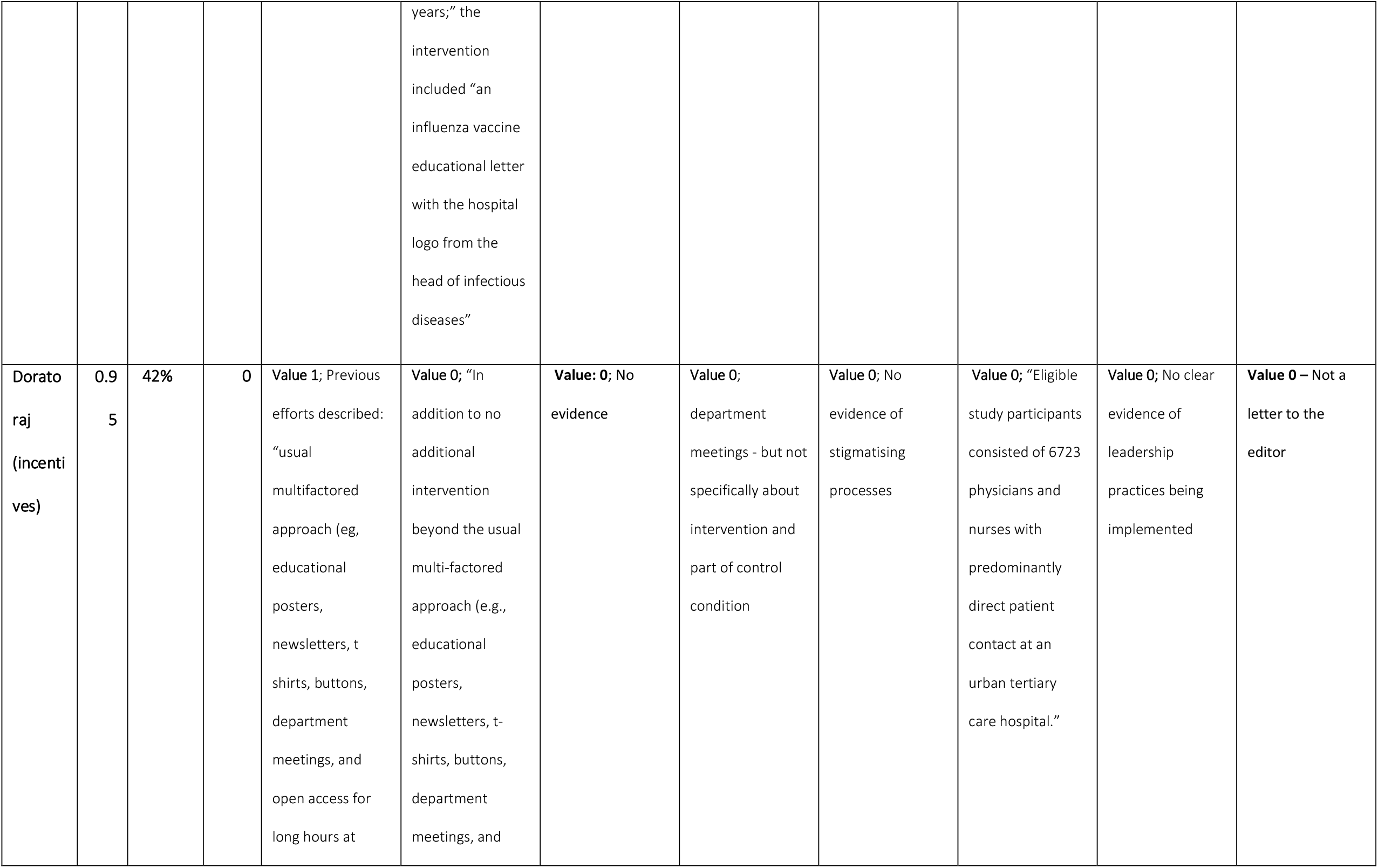
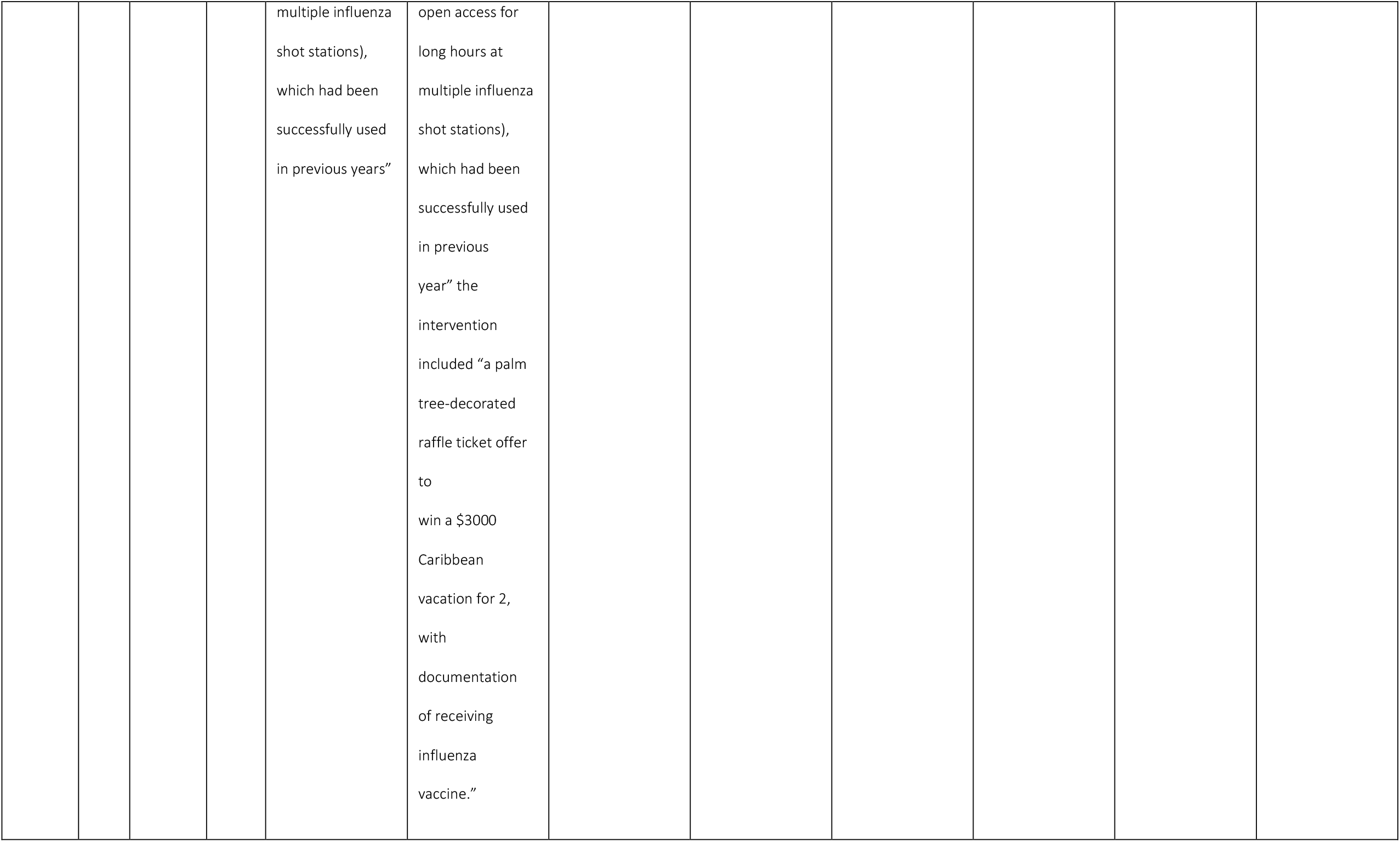
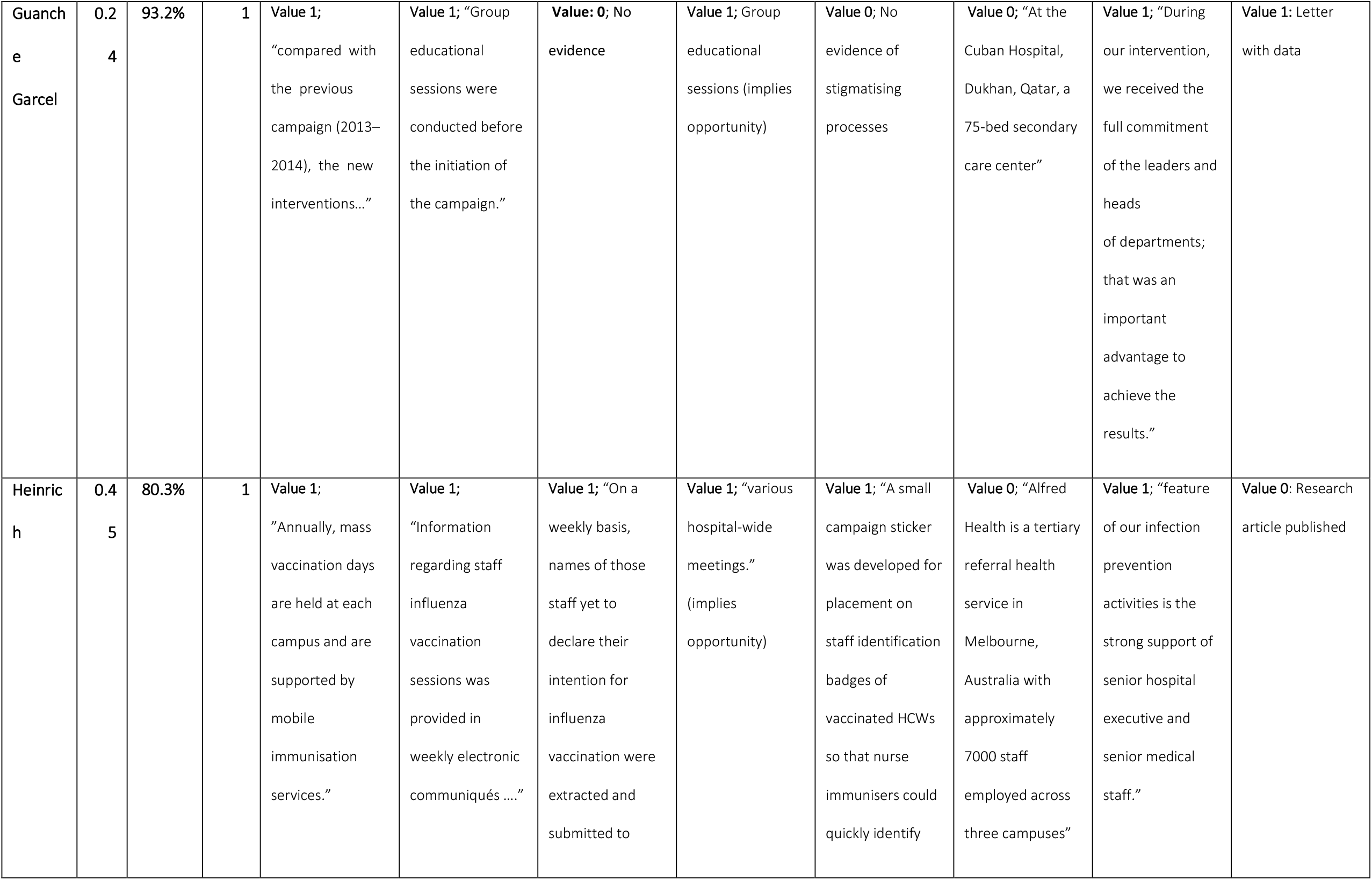
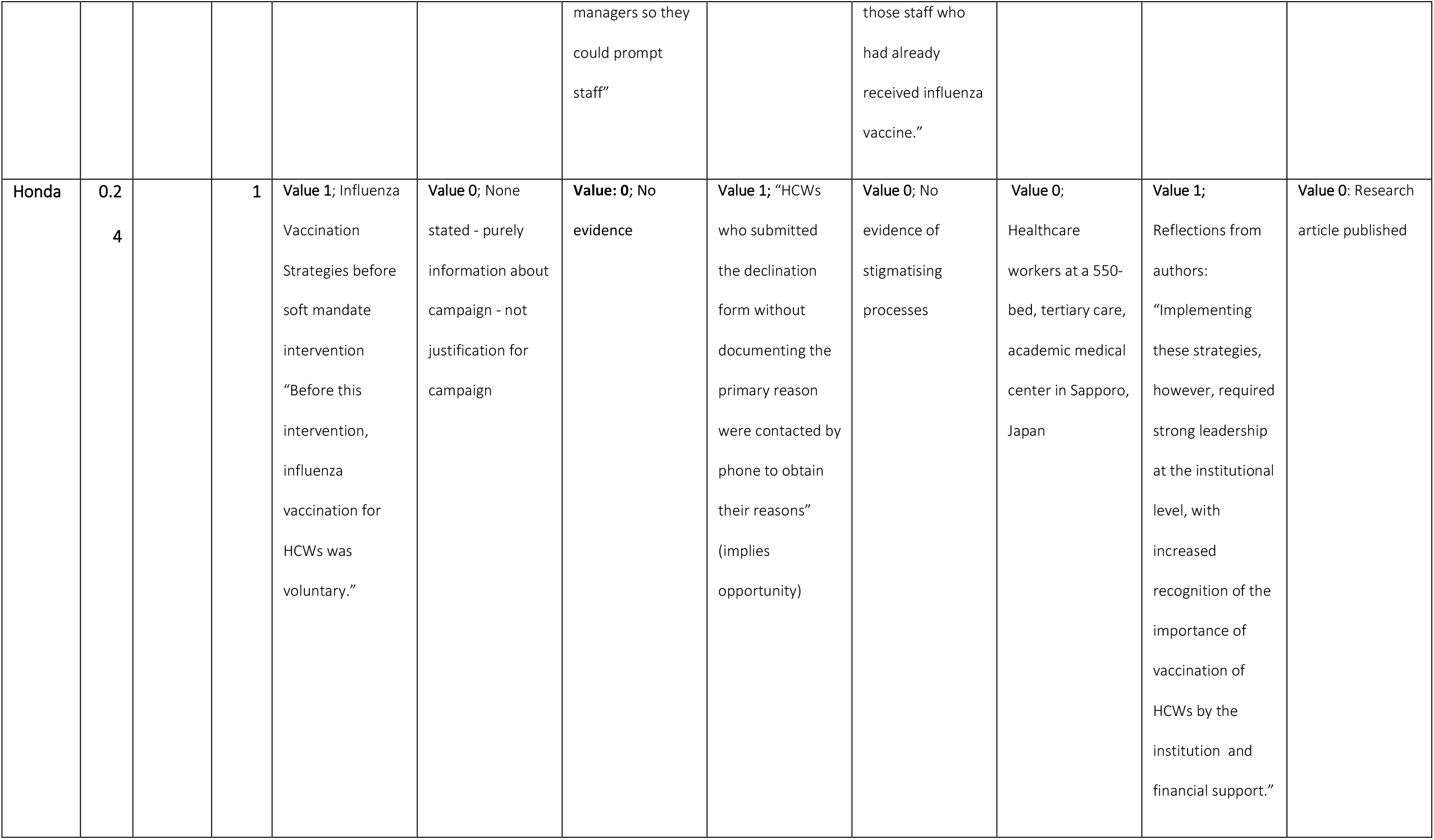
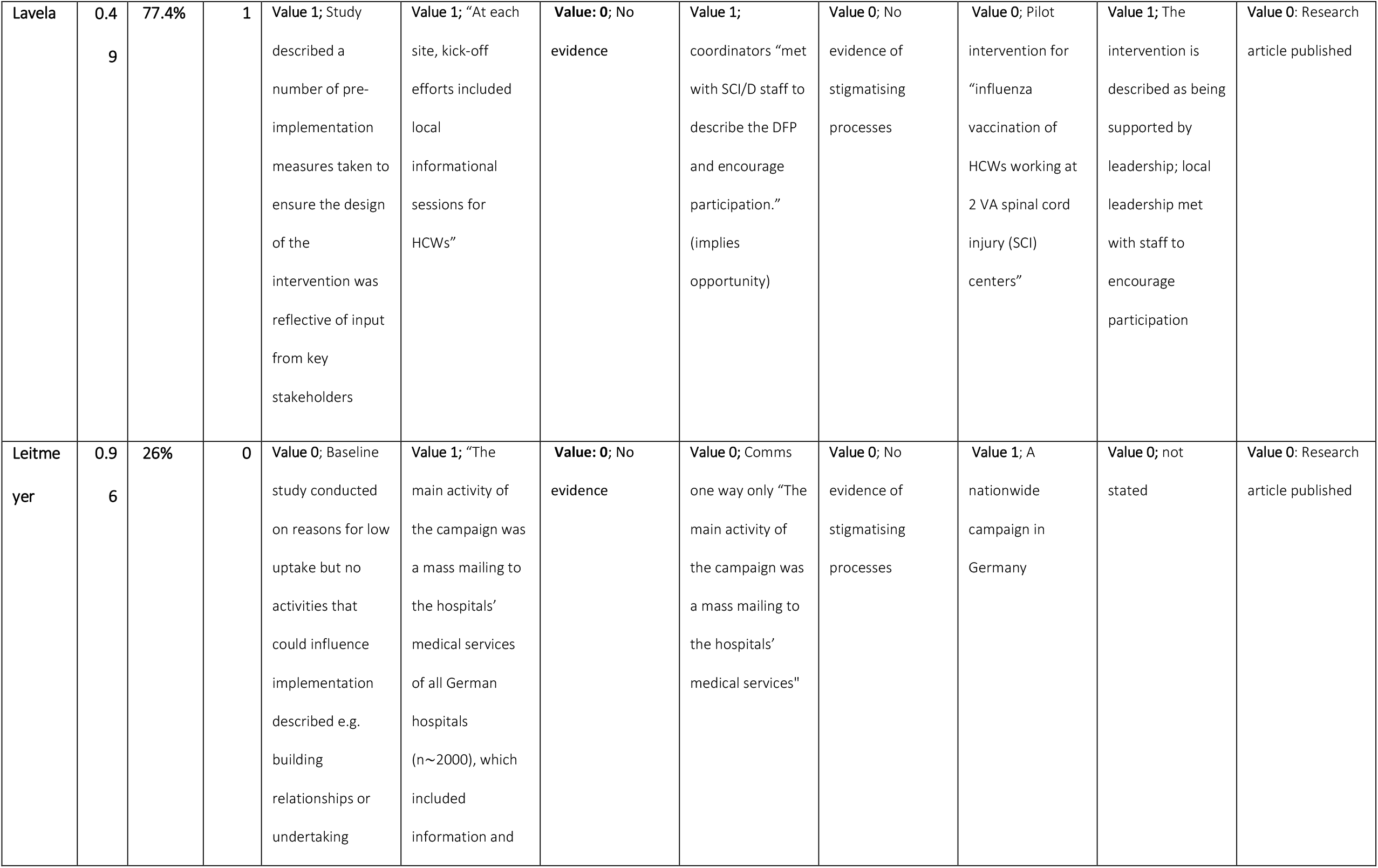
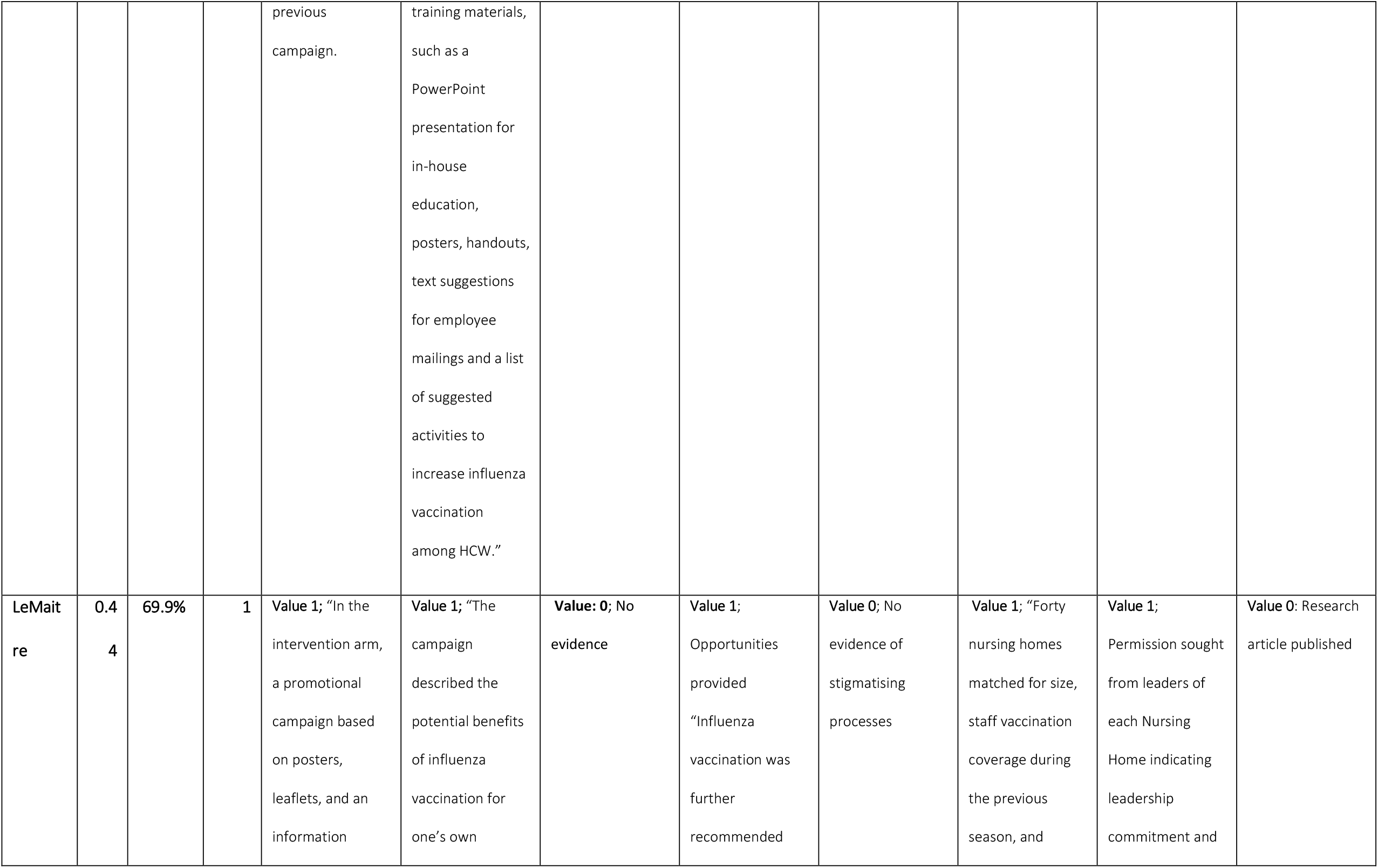
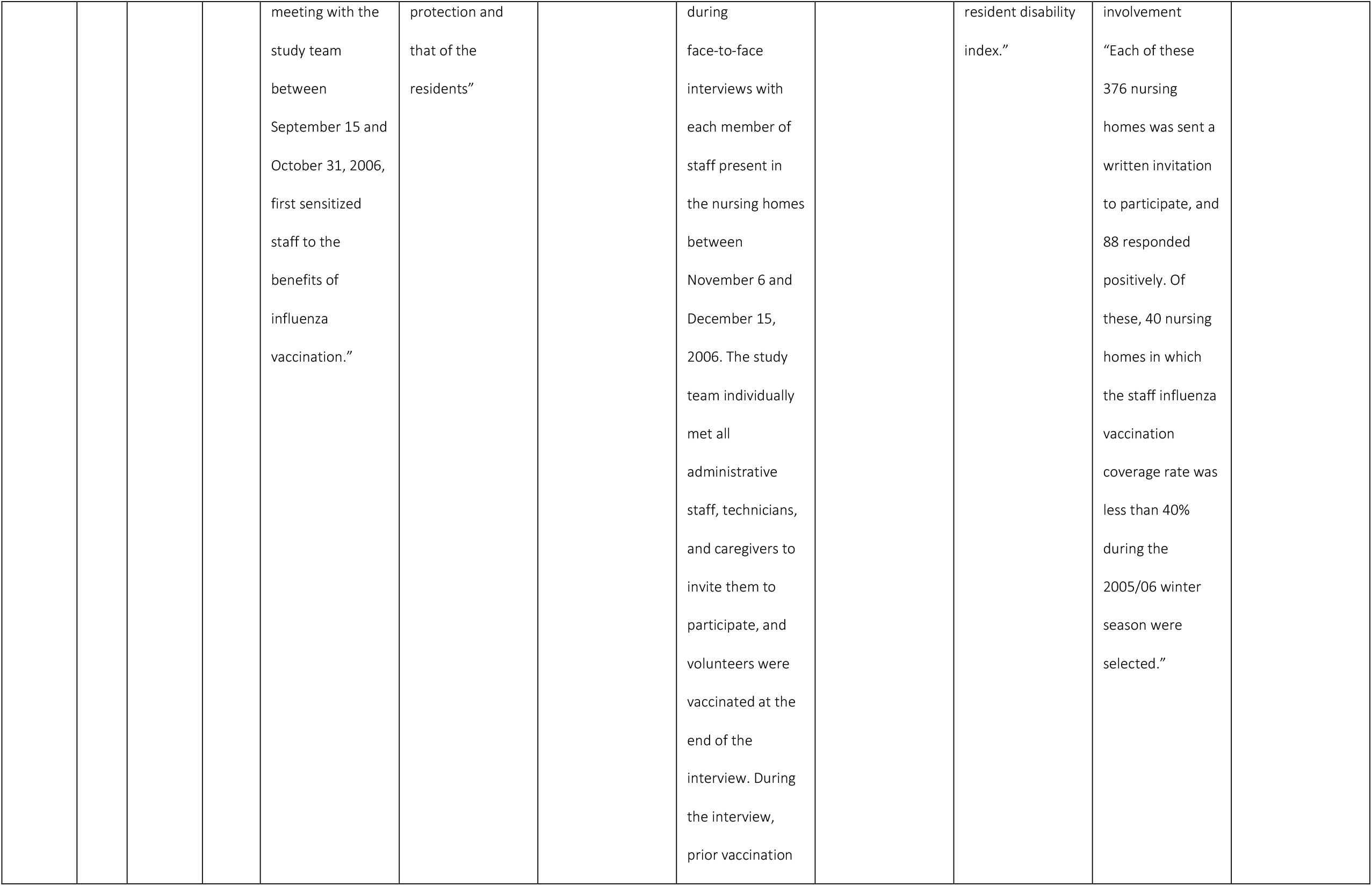
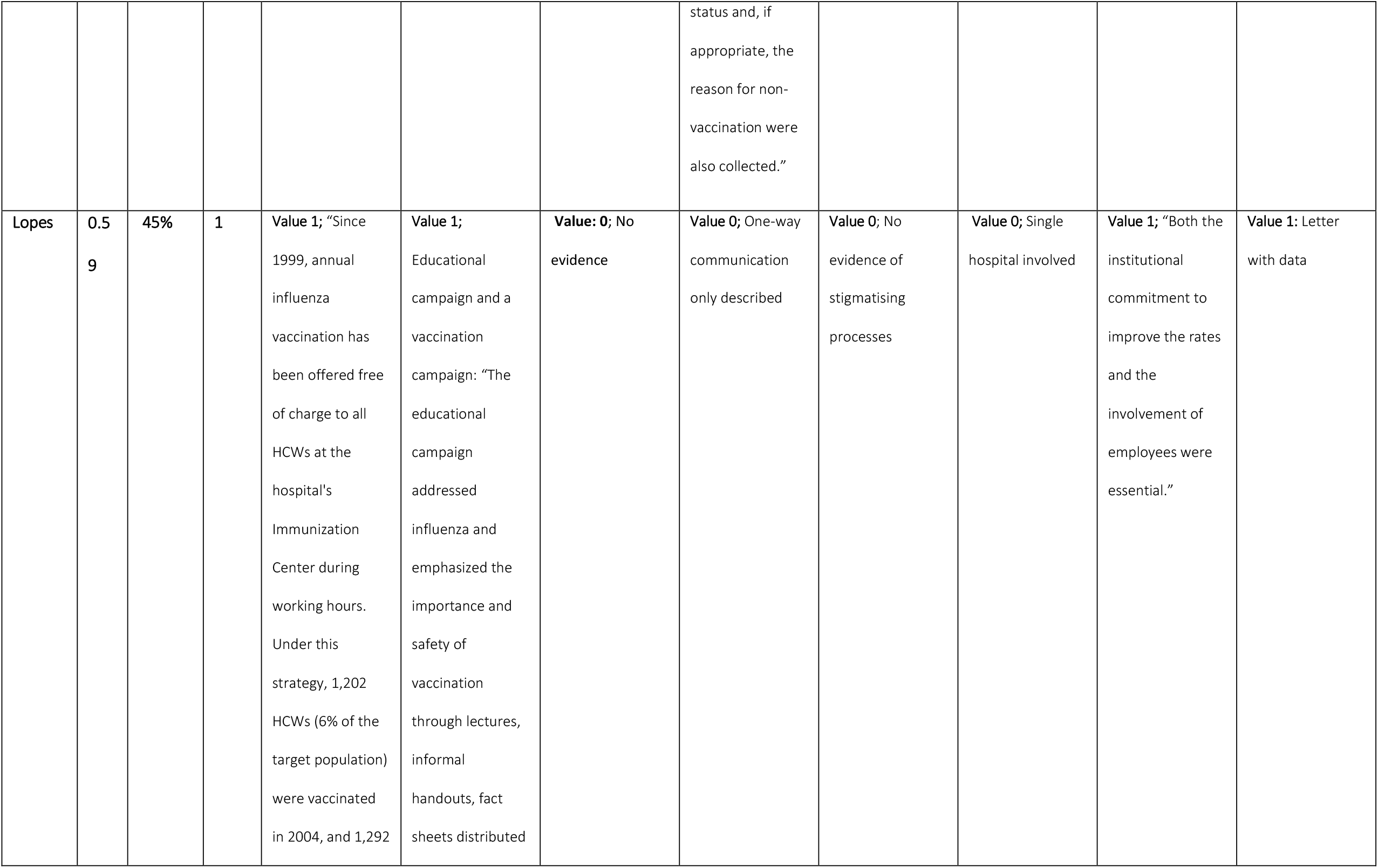
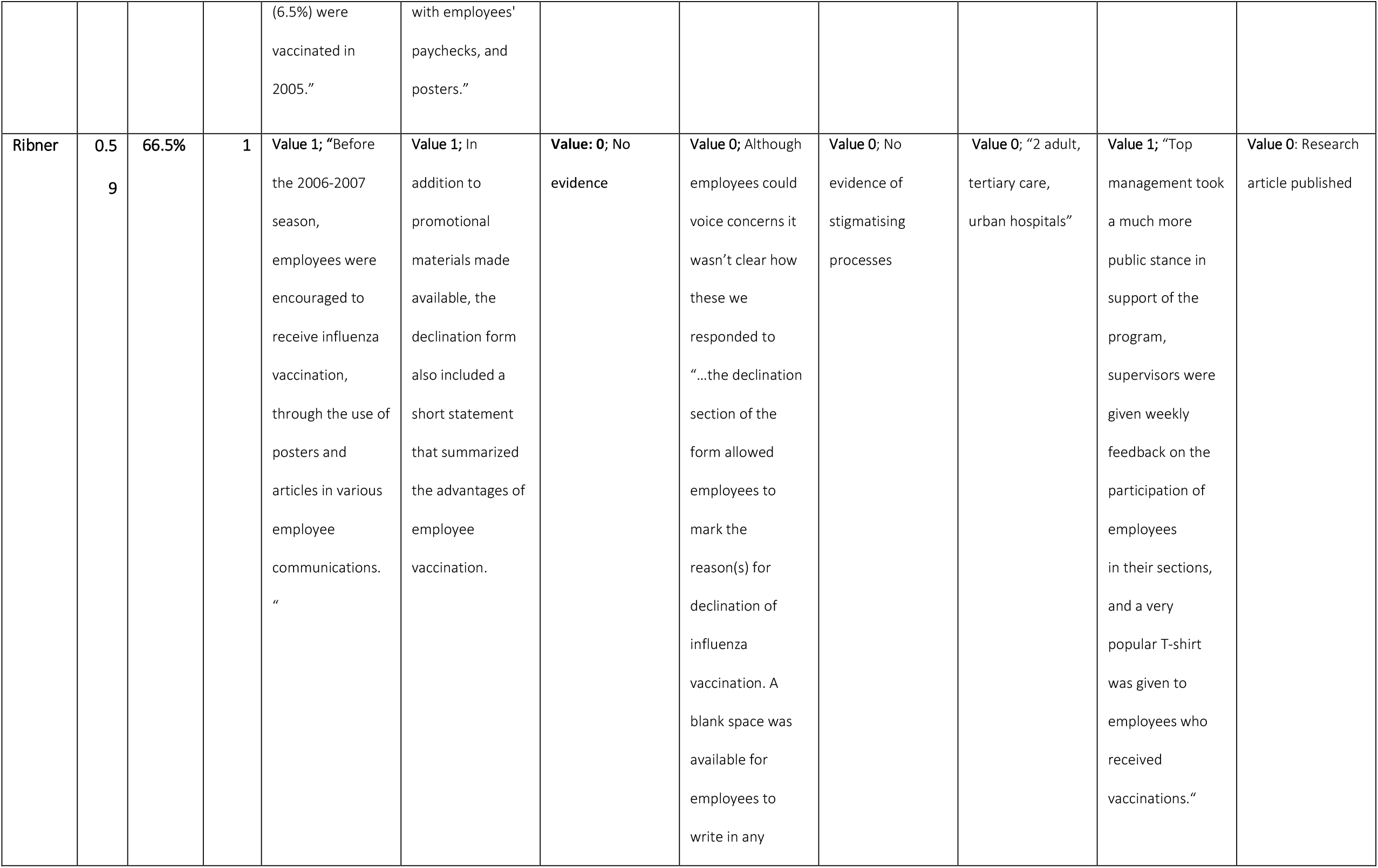
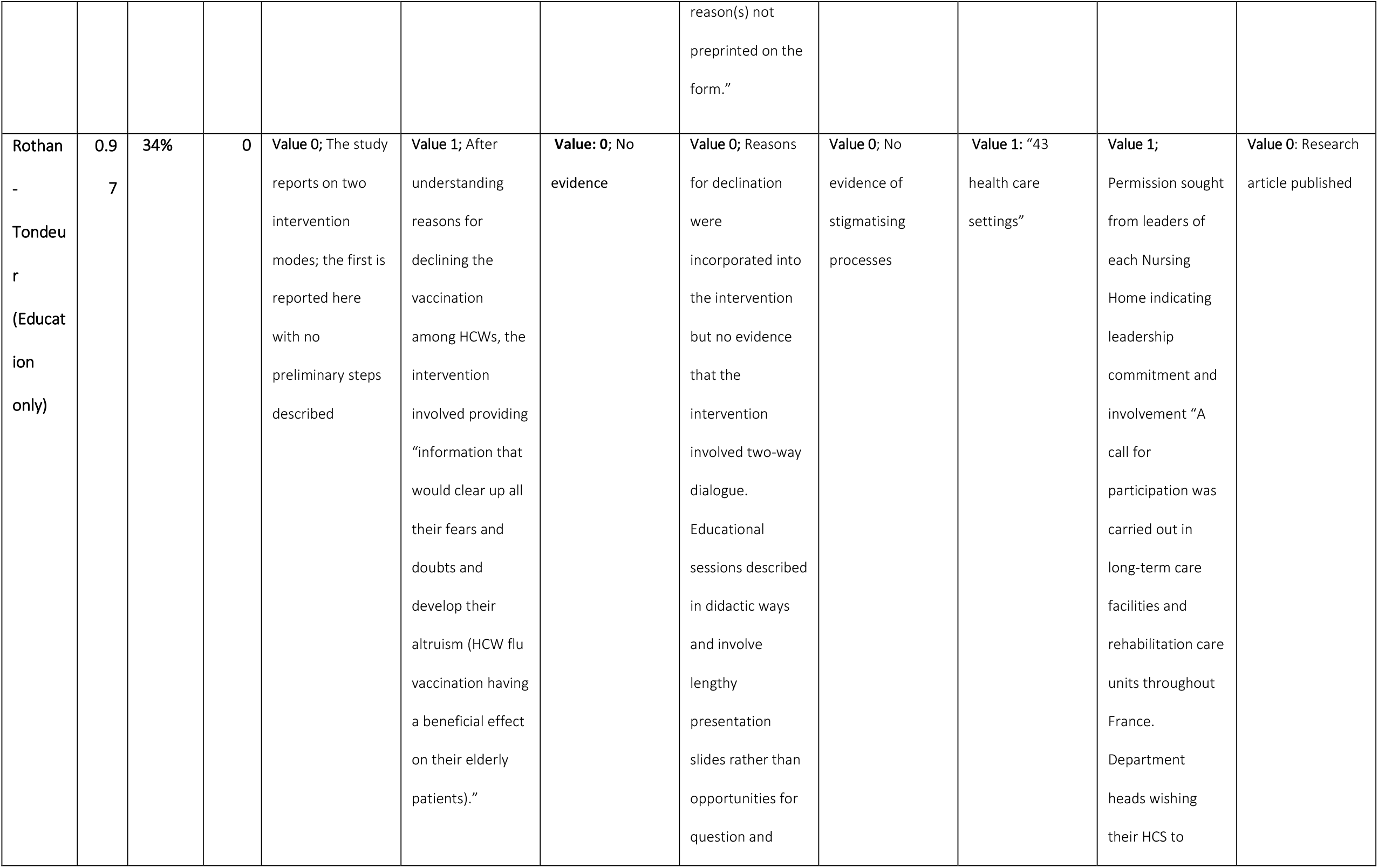
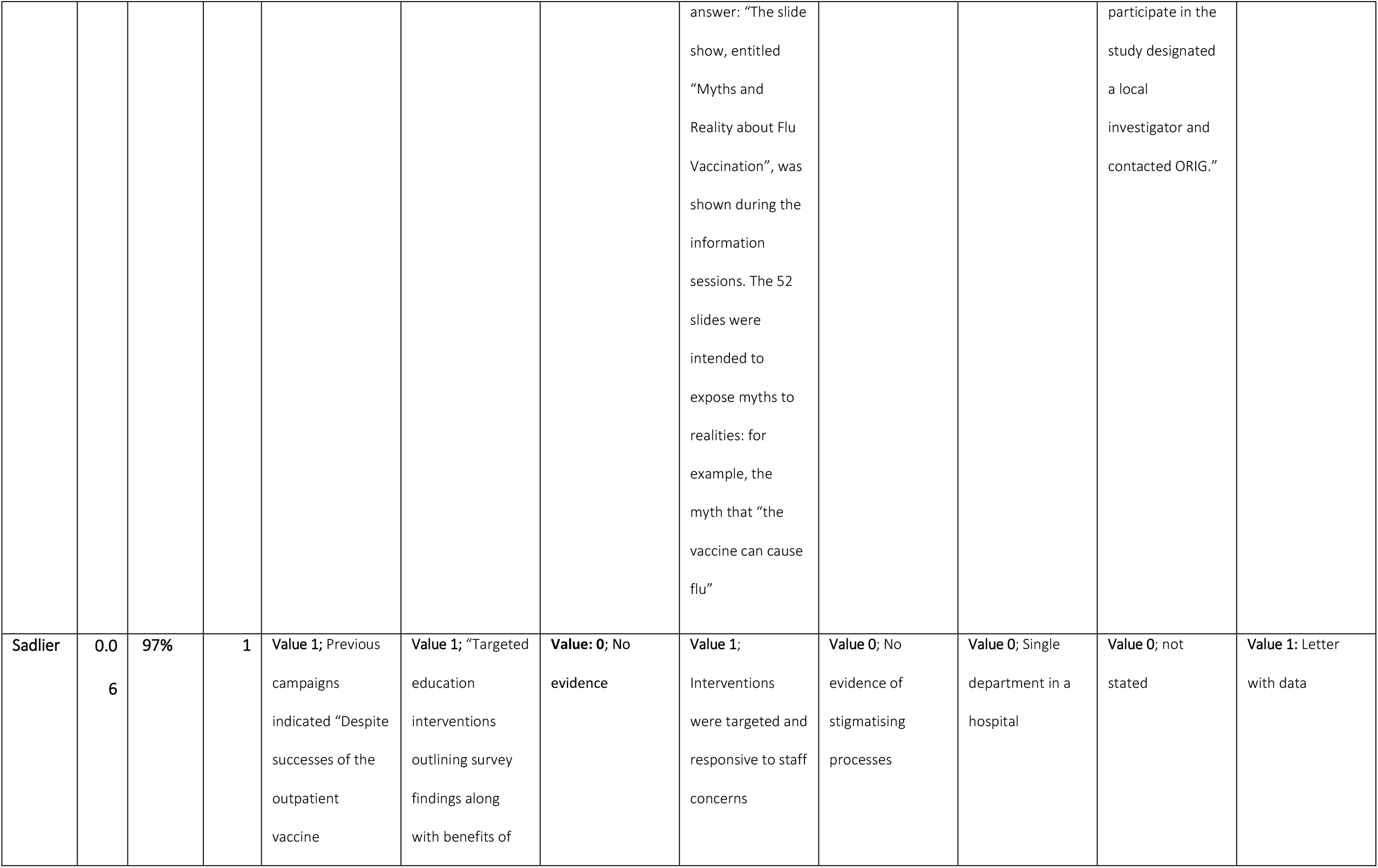
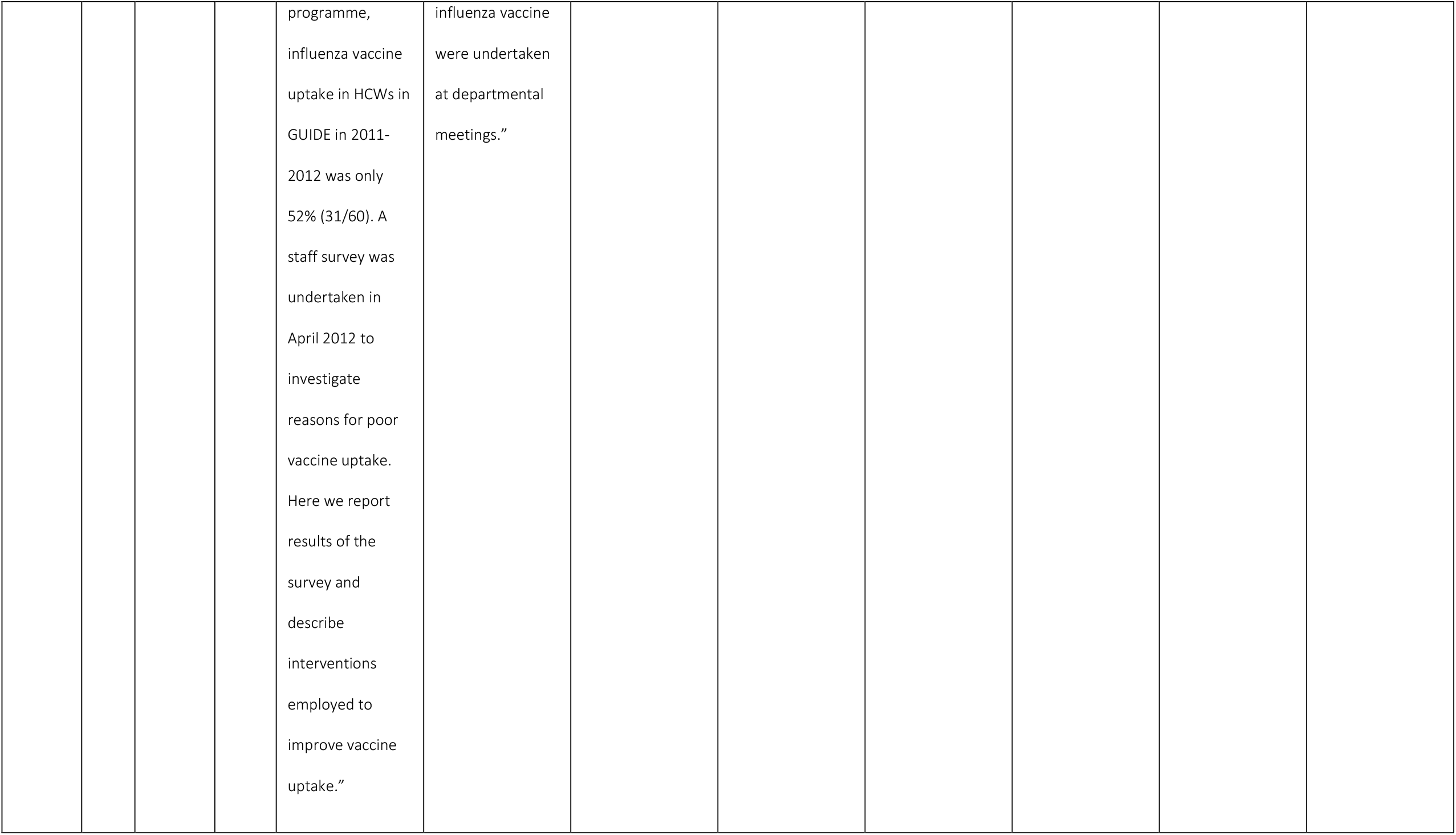
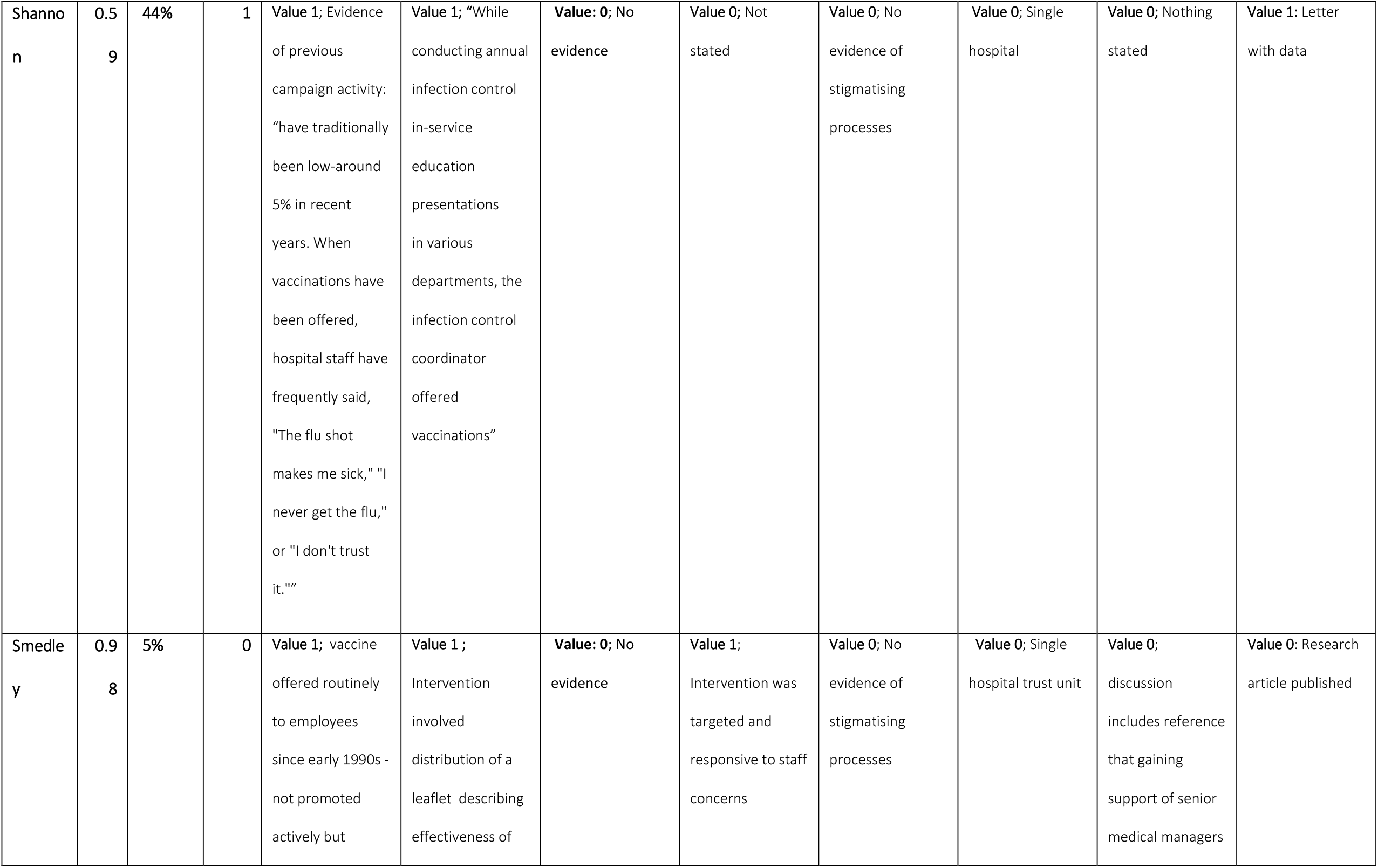
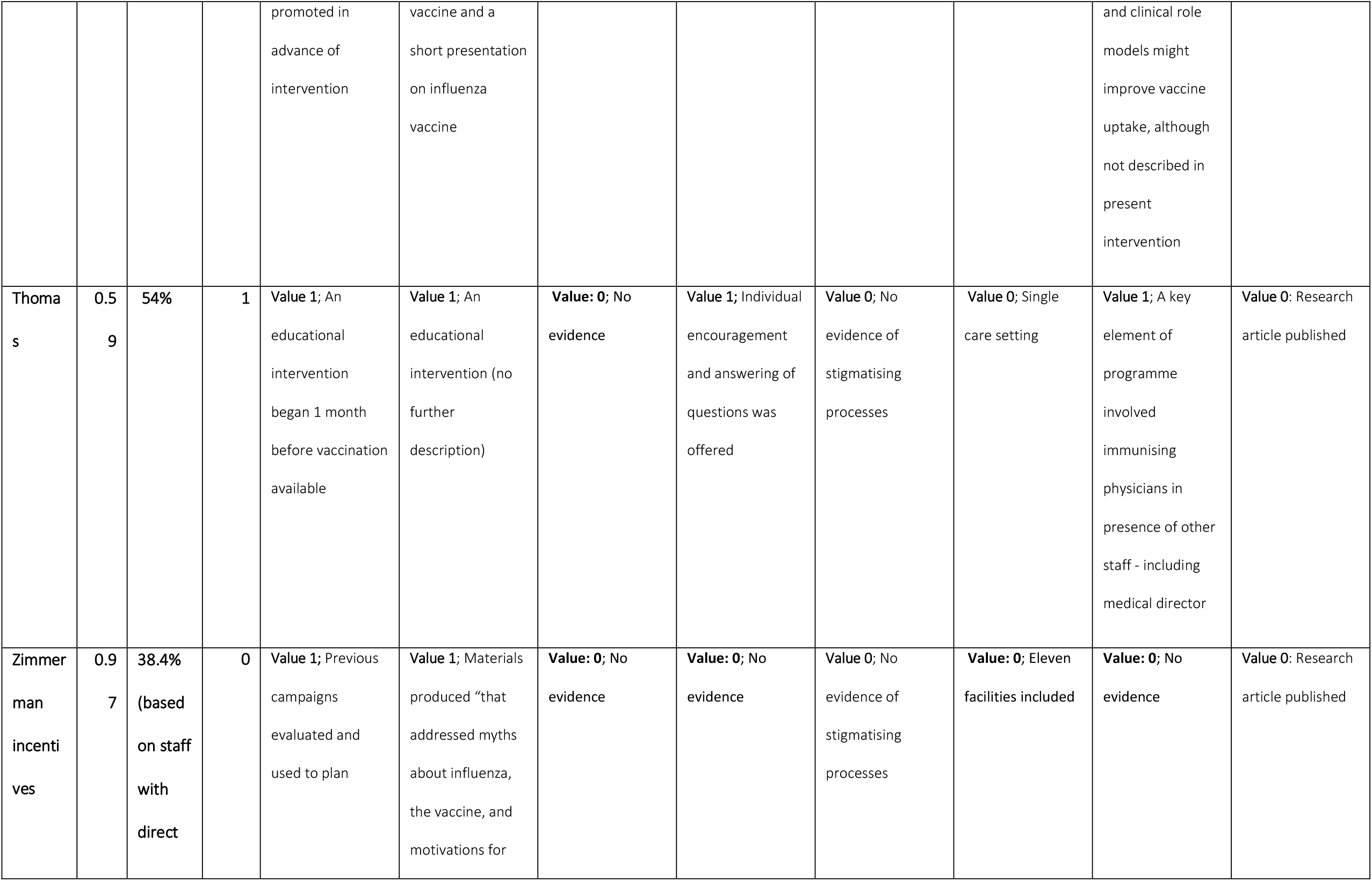
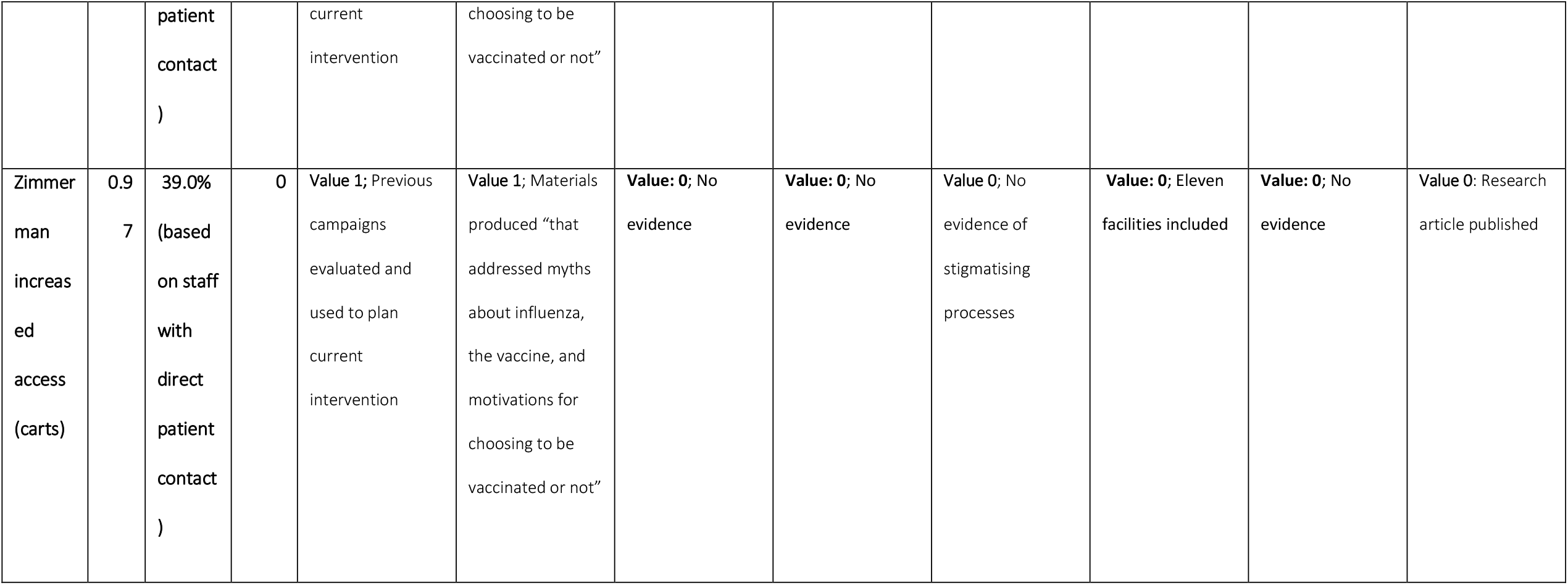

